# Efficacy inference in early-phase non-controlled clinical trials via Bayesian biomarker deconvolution

**DOI:** 10.64898/2026.06.26.26356652

**Authors:** Chris Humphries, Alastair M Kilpatrick, Jennifer A Cartwright, Charlotte Potter, Anuruddika J Fernando, Maria Elena Candela, Janet Man, Rhona Aird, Kenneth J Simpson, Marcus J Lyall, Philip Starkey Lewis, Aryelly Rodriguez, Christopher J Weir, James W Dear, Stuart J Forbes, Linus J Schumacher

## Abstract

Early-phase clinical trials of therapies for acute organ injury are typically small, uncontrolled, and must infer treatment activity using only tissue-damage biomarker changes over time. Interpretation is confounded by the temporal overlap of ongoing tissue damage and biomarker clearance. We address this with a Bayesian deconvolution framework that fits individual patient biomarker trajectories with an exponentially-modified Gaussian model. This model separates injury kinetics (peak release rate, injury duration, time to peak) from biomarker clearance, using a historic cohort as a null distribution. In a simulated phase 1 dose-escalation regenerative therapy trial, the framework reduces the minimum detectable treatment effect from 67.5% to 24.5% (a 2.76-fold improvement) and supports dose selection. Applied to published case-series data for another therapeutic, the framework recovers per-patient pharmacodynamic signatures consistent with pre-clinical mechanistic studies and supports smaller prospective clinical trial design. With indication-specific recalibration, the framework architecture conceptually transfers to other acute organ injuries where serum biomarkers reflect tissue damage. This offers a route to significantly reducing clinical trial sizes, supporting therapy dose-finding in non-controlled clinical trials, and identifying pharmacodynamic signatures.

## Introduction

The US FDA’s January 2026 draft guidance on Bayesian methodology in clinical trials formally endorses Bayesian approaches for dose-finding and the construction of informative priors from historical data.(1) This creates a regulatory pathway for Bayesian frameworks that extract pharmacodynamic signals from small, uncontrolled early-phase trials. To our knowledge no established methodology operationalises this opportunity for acute organ injury, where trials enrol acutely ill patients whose disease is evolving at the time of treatment.(2,3)

Tissue-specific biomarkers are routinely used as proxies for tissue damage in trials and clinical practice, and are often the only objective measure of therapeutic effect at the target organ.(4) The central analytical challenge is that any serum tissue-damage biomarker reflects the superposition of ongoing injury and clearance.(5) This is an interaction seen across acute organ injury (myocardial infarction, rhabdomyolysis, pancreatitis, neurological injury)(6–12); naive decay models mis-estimate clearance during the period of temporal overlap, which is precisely when patients in acute-injury trials are typically treated (Fig.1a, Supplementary Fig.1).(13)

**Figure 1.**
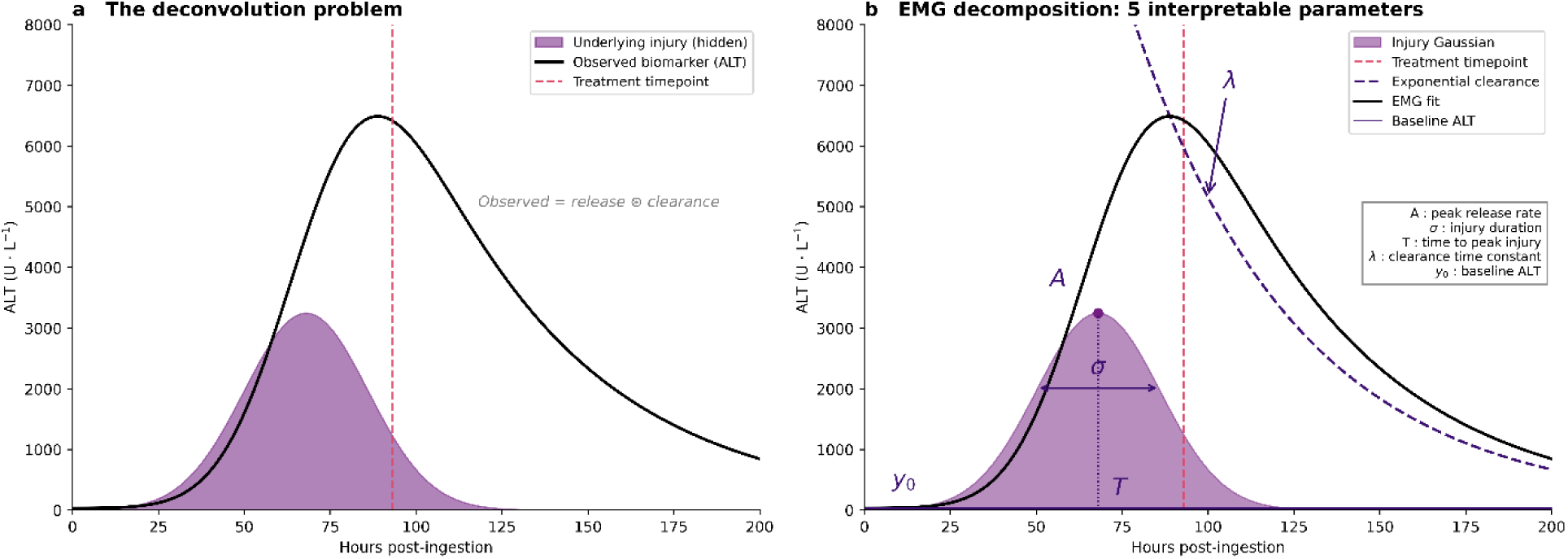
Framework overview and EMG model. (a) The deconvolution problem: serum biomarkers in acute organ injury reflect the superposition of injury-driven biomarker release (shaded area) and first-order clearance. This confounds naive comparison of observed trajectories when candidate treatments are administered while injury is ongoing. (b) The exponentially-modified Gaussian (EMG) decomposition: the observed ALT trajectory is the convolution of a Gaussian injury process with first-order clearance, yielding five structural parameters: peak release rate (A), injury duration (σ), time to peak injury (T), clearance time constant (λ), and baseline ALT (y₀). In our model, y₀ is fixed at 30 U·L⁻¹ (mid-range of normal limits) and other parameters are inferred per patient. Each fit additionally infers the patient’s ingestion time and a measurement-noise scale.

A second challenge is that early-phase trial designs are uncontrolled, with small patient numbers spread across dose cohorts.(14) Phase 1 trials of regenerative cell therapies in this setting typically nominate a biomarker-based activity analysis, such as isotonic regression, across dose groups.(15) This is defensible given the trial limitations but the test is constrained: the apparent effect is confounded by treatment timing relative to injury, it tests a monotonic trend across treated patients only, and it does not adjust for the pre-treatment individual variation in clearance rate, injury size and treatment timing characteristic of acute-injury populations.(16)

Our central proposition is that this constrained inferential setting can be substantially improved by replacing the raw biomarker outcome with individual counterfactual predictions (each patient’s expected untreated trajectory), conditioned on each patient’s own pre-treatment kinetic parameters (Fig.1b). We develop the framework in paracetamol-induced acute liver injury (APAP-DILI; the most common cause of acute liver failure in many countries).(2) In APAP-DILI, aminotransferases such as alanine aminotransferase (ALT) act as the typical biomarker proxy for injury severity.(17) These biomarkers enter the circulation as hepatocytes undergo necrosis, and are cleared by first-order kinetics primarily by liver sinusoidal endothelial cells (LSECs) and resident hepatic macrophages.(10–12,17–19) LSECs and resident macrophages are themselves injured in APAP-DILI, and more severe injury is associated with prolonged aminotransferase clearance.(19–22) An intervention that restored endogenous clearance capacity or truncated ongoing hepatocyte injury would alter the corresponding kinetic parameter.

Here, we develop a framework (combining per-patient exponentially-modified Gaussian biomarker deconvolution, functional principal component analysis and a historic-cohort null) that converts each patient’s routinely sampled ALT trajectory to generate counterfactual predictions, against which the post-treatment trajectory can be assessed. The framework lowers the minimum effect size detectable in small uncontrolled dose-escalation designs and separates injury kinetics from clearance kinetics (supporting mechanism-targeted readouts that support distinguishing a regenerative therapy from a therapy which prevents injury). The framework also produces per-patient posteriors on each kinetic parameter to support continuous dose-decision-making and accommodates sparse and irregularly sampled trajectories. The architecture is demonstrated in APAP-DILI, but transfers conceptually to any acute organ injury where serum biomarkers reflect tissue damage, subject to adjustments for injury kinetics and refitting with appropriate priors.

## Results

### Cumulative tissue-damage biomarker exposure is a proxy for tissue injury

To validate our foundational assumption that cumulative biomarker exposure reflects tissue injury, we used a murine model of paracetamol hepatotoxicity(n=44 paracetamol-treated mice; Supplementary Methods 1, Supplementary Figures 2 and 3, and Supplementary Table 1). The integrated ALT area-under-curve tracked the percentage of centrilobular necrosis quantitatively across the time course (Supplementary Figures 2 and 4), supporting use of cumulative ALT exposure as a tissue-injury proxy in the preclinical setting and motivating its decomposition into separable injury and clearance components in human data.

We compared four feature-extraction approaches applied to serial ALT measurements against histological necrosis at each timepoint (Supplementary Table 2). Point measures and peak-anchored measures degraded as the interval from injury increased, while cumulative ALT exposure (AUC to cull; the area under the ALT–time curve up to the cull timepoint) maintained strong correlation with necrosis across all timepoints (Supplementary Table 2), reflecting the increasing influence of clearance kinetics on point measurements.

An ordinary least-squares log-log regression on all 44 mice (necrosis=θ_1_ × time^{θ_2_} × ALT AUC; both θ p<0.001, n=44, through-origin R^2^=0.91; Fig.2a-b; Supplementary Analysis 1) confirmed ALT AUC as a quantitative surrogate for necrosis when time since injury is accounted for (RMSE 0.114, bias<0.01, 95% limits of agreement −0.22 to 0.23; Supplementary Fig.5).

**Figure 2.**
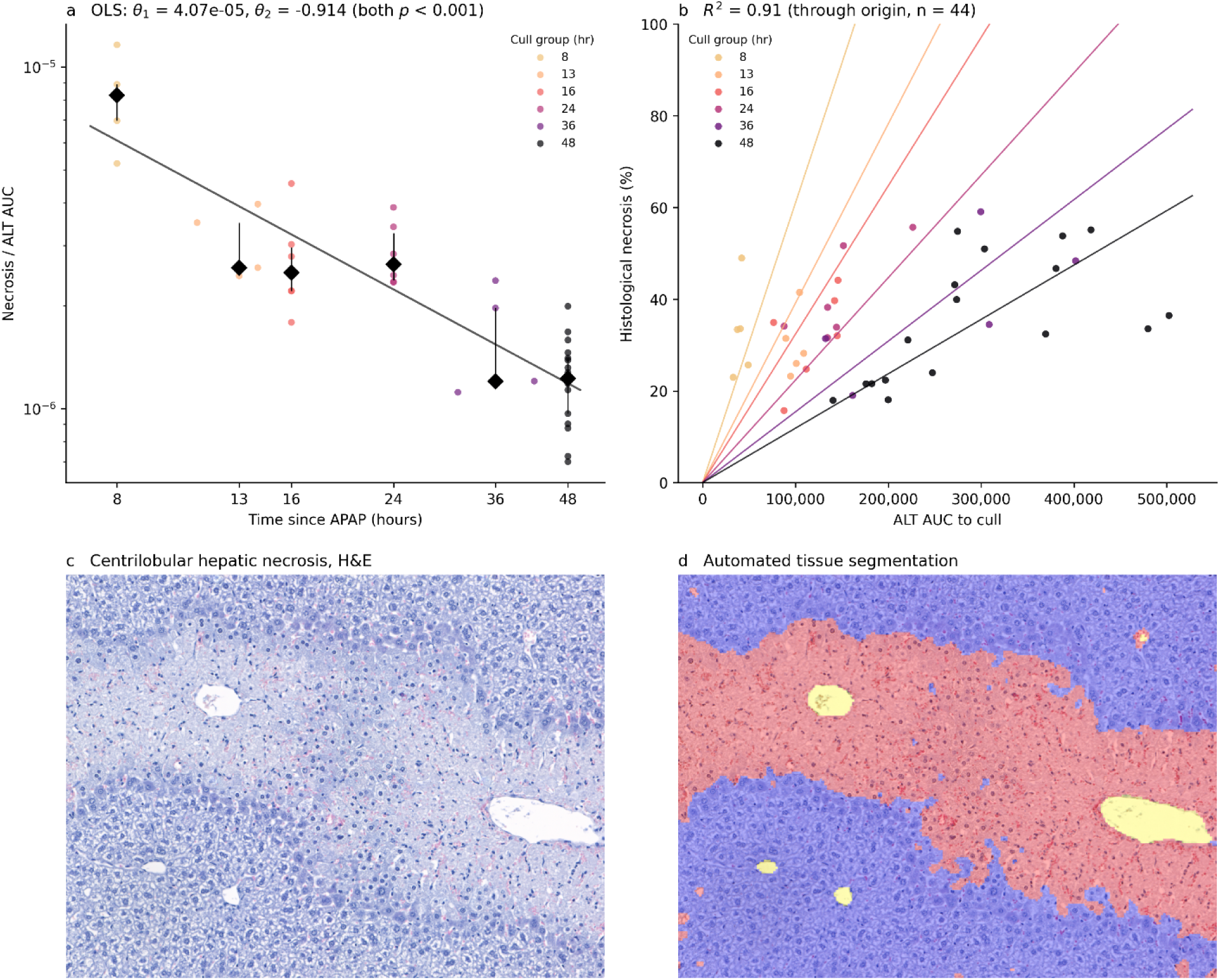
Preclinical validation of ALT AUC as a necrosis surrogate. (a) The ratio of histological necrosis to cumulative ALT AUC for each of 44 APAP-treated mice follows a power-decay function over time. The OLS fit (grey line, θ_1_=4.07 × 10^-5^, θ_2_=-0.914, both p<0.001) is overlaid on individual mice (small points, coloured by cull group) and group medians (diamonds) plus or minus interquartile range. (b) Equivalent log-log linear regression of ALT AUC and time against necrosis for all 44 mice, back-transformed to the original linear scale, demonstrating an uncentred R^2^=0.91 (through origin) across cull groups. (c) Representative centrilobular hepatic necrosis on haematoxylin and eosin (H&E) staining. (d) Automated tissue segmentation of the same field, distinguishing necrotic from viable parenchyma.

The time correction revealed a limitation of raw biomarker values: using naïve AUC conflates injury-driven ALT release with the clearance tail, and the empirical power-decay can only correct for this globally, without decomposing the two processes. A model that explicitly deconvolved injury from clearance would replace this empirical correction with a mechanistic decomposition, while yielding interpretable parameters (injury amplitude, duration, timing, and clearance rate) as pharmacodynamic endpoints - and would support inference on either side of the injury-clearance convolution.

### An exponentially modified Gaussian model deconvolves injury and clearance in human ALT trajectories

Using a cohort of 195 human patients admitted for the treatment of paracetamol overdose, we modelled each patient’s ALT trajectory with a Bayesian exponentially-modified Gaussian (EMG) model that separates injury kinetics from first-order clearance. Each patient yielded posterior estimates of the EMG shape parameters: peak release rate (A), injury duration (σ), time to peak (T), clearance time constant (λ), with baseline (y₀) fixed at 30 U·L⁻¹ as well as a latent ingestion time and inferred measurement-noise scale (Fig.1).

By separating injury and clearance processes, the EMG model parameters become biologically interpretable. The EMG arises as the convolution of a Gaussian injury process with a first-order clearance mechanism, fitted under a single-injury-event assumption per patient. Full model specification, priors and convergence diagnostics are supplied in Supplementary Methods 2.

We fitted the EMG model within a Bayesian inference framework to 195 patients with APAP-DILI and peak observed ALT>1000U/L and at least three serial ALT measurements, from NHS Lothian hospitals (Supplementary Figures 6 and 7, Supplementary Table 3). All patients received N-acetylcysteine (NAC) as standard of care. The full dataset comprised 2,036 ALT observations (median 9 per patient, range 3 to 35). Of these 195 patients, 183 had post-peak data allowing their inclusion in functional principal component analysis (fPCA); patients with sparse or divergent data are characterised under Supplementary Table 6, and 130 had sufficient post-peak data to simulate treatment effects for the power analysis.

The fitted model produced individual posterior estimates of all five EMG parameters fitted to each patient’s complete ALT trajectory under weakly-informative, literature-based priors (Supplementary Methods 2). A sensitivity analysis using 10-fold cross-validation to derive cohort-informed priors did not improve predictive accuracy, so the full-data fit with the pre-specified priors was used throughout (Supplementary Table 4).

For each patient, the fitted EMG model generates a counterfactual prediction of the ALT trajectory expected in the absence of intervention, using only pre-treatment observations (Fig.3). Any systematic departure of the observed post-treatment trajectory from this prediction implies a change in the underlying injury or clearance dynamics.

**Figure 3.**
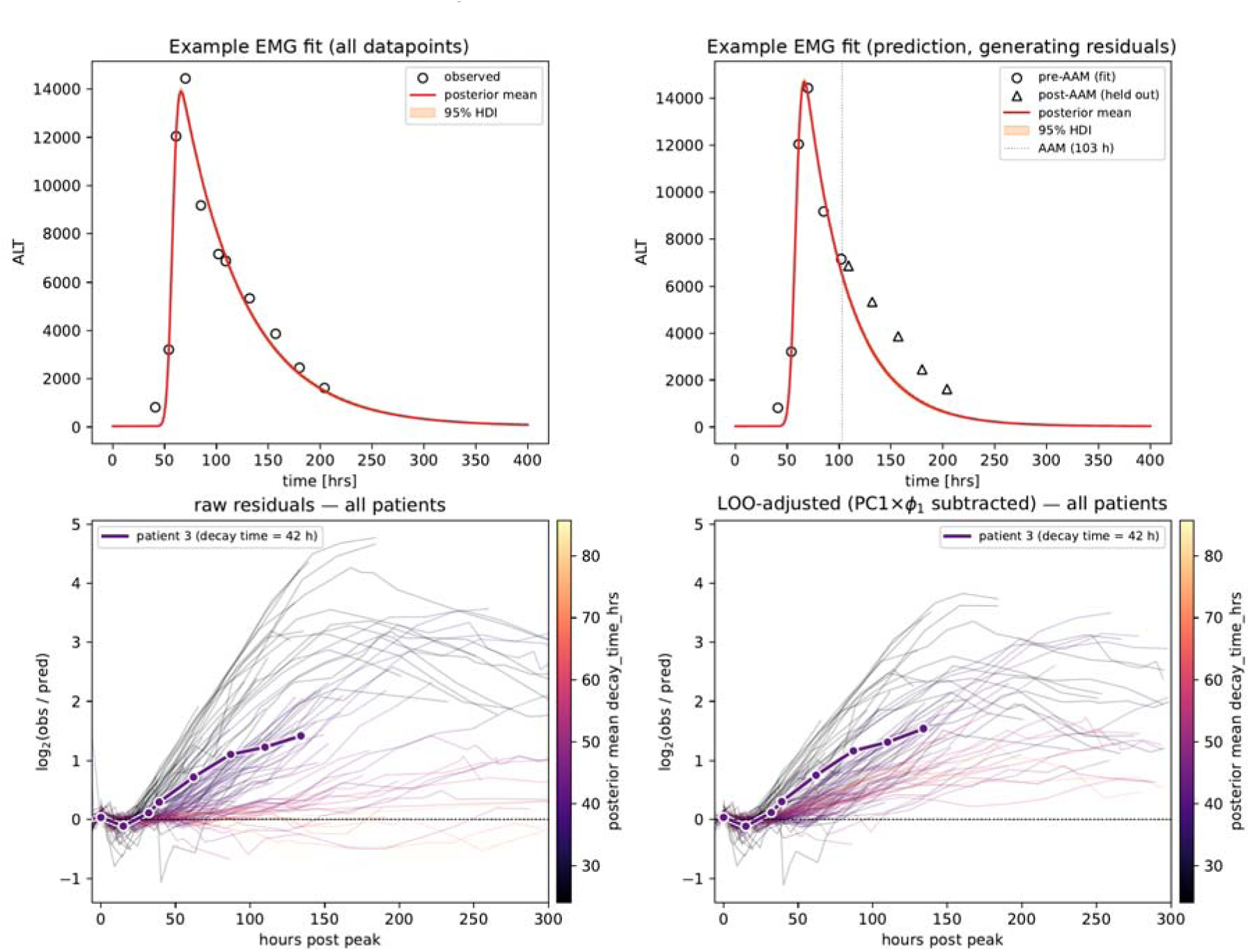
EMG model fits, counterfactual prediction, and population-level residuals. **(a)** Posterior EMG fit (red line, 95% credible interval shading) to all observations (black points) for a representative patient. Additional representative fits are in Supplementary Fig.8. **(b)** Counterfactual posterior predictive for the same patient: the EMG is fit to pre-intervention-point observations only (black circles; assessment point at approximately 50% of post-peak amplitude), then extrapolated forward (red line, 95% CrI shading). Predicted observations demonstrate non-zero residuals, which are captured across the cohort to quantify the predictive error structure. **(c)** Raw residual trajectories for the population (one line per patient), coloured by clearance time constant λ. The pronounced λ-dependent autocorrelation motivates leave-one-out (LOO) regression adjustment. **(d)** After subtracting the LOO-regression-predicted Principal Component 1 (PC1) contribution at each timepoint, trajectories can be treated as a single population in downstream testing.

Validation of the the EMG model used both preclinical and clinical anchors. In the murine model, integrated ALT exposure correlated with the percentage of centrilobular necrosis on histology (Fig.2). In humans, where biopsy is not feasible, the EMG-derived Gaussian injury AUC correlated with peak international normalised ratio and peak bilirubin - established biomarkers of liver function (Supplementary Analysis 2).(2,15,17)

### Functional Principal Component Analysis and Leave-One-Out regression convert per-patient residuals into a population-scale treatment score

To allow interrogation of sparse, irregularly-sampled longitudinal data, we applied PACE (Principal Analysis by Conditional Expectation), a functional PCA method designed specifically for this scenario (Supplementary Methods 3).(23) Each patient’s (log2) EMG residuals (observed/predicted) at each ALT observation were projected into a reduced dimensional space using PACE-fPCA. The first three PCs explained 95% of the variance in residual trajectories (Fig.4a-b); PC1 explained 86.3% and carries the dominant clearance-modification signal. We applied leave-one-out (LOO) regression on the clearance time [log(λ)]; the resulting PC1_adj_ score is, by construction, uncorrelated with each patient’s pre-treatment kinetics, allowing the historic cohort to serve as a null distribution against which post-treatment trajectories can be scored without requiring concurrent controls (Supplementary Methods 3 and 4).

**Figure 4.**
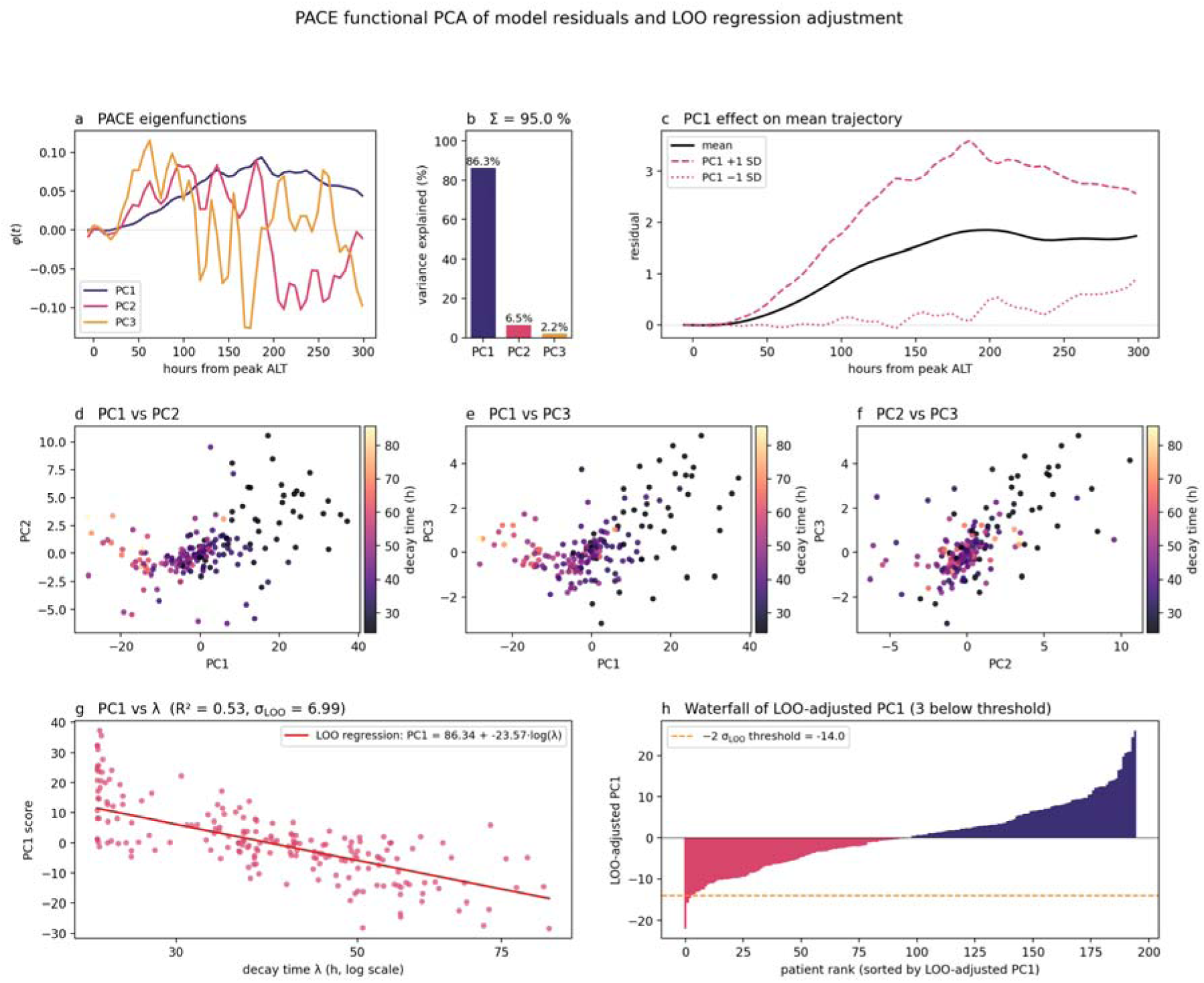
Functional PCA of model residuals and LOO regression adjustment. **(a)** The three PACE eigenfunctions capturing the dominant modes of residual variation in the post-peak window, **(b)** accounting for 95.0% of the total variance. **(c)** The effect of PC1 on the mean residual trajectory: plus or minus 1 SD of PC1 corresponds to a sustained shift, consistent with prolonged or accelerated clearance. **(d-f)** Pairwise scatter plots of the per-patient PC scores, showing that PC1 carries most of the λ-dependent signal whereas PC2 and PC3 reflect higher-order shape differences. **(g)** PC1 versus log(λ): the empirical LOO regression line is overlaid; residuals from this regression define PC1_adj_. **(h)** Waterfall plot of LOO-adjusted PC1 scores.

In combination, this pipeline improves treatment-effect detection sensitivity. PC1_adj_ is computed by fitting an EMG to each patient’s pre-treatment data (absorbing baseline severity, peak ALT, peak timing), projecting the post-treatment residual onto PACE eigenfunctions, and subtracting the LOO-predicted PC1 contribution given the patient’s own λ. The result enters the slope test as a within-patient counterfactual deviation rather than a raw outcome. Each patient’s score is computed from their available ALT measurements via conditional expectation, without requiring observations at protocol-specified timepoints. Patients whose sampling deviates from the planned schedule remain scorable, and the same property supports retrospective application to routine clinical biomarker data.

### Semi-synthetic validation confirms the framework’s treatment-effect detection sensitivity across patient phenotypes

To test the framework’s detection sensitivity across phenotypes, we overlaid 5 simulated clearance-acceleration effects onto three held-out patients (not in the null cohort) spanning the null cohort’s full clearance range and examined whether PC1_adj_ recovered the imposed effect monotonically. PC1_adj_ shifted monotonically with treatment strength at each end of the clearance range (Fig.5), supporting use across the heterogeneous patient populations seen in phase 1 trials.

**Figure 5.**
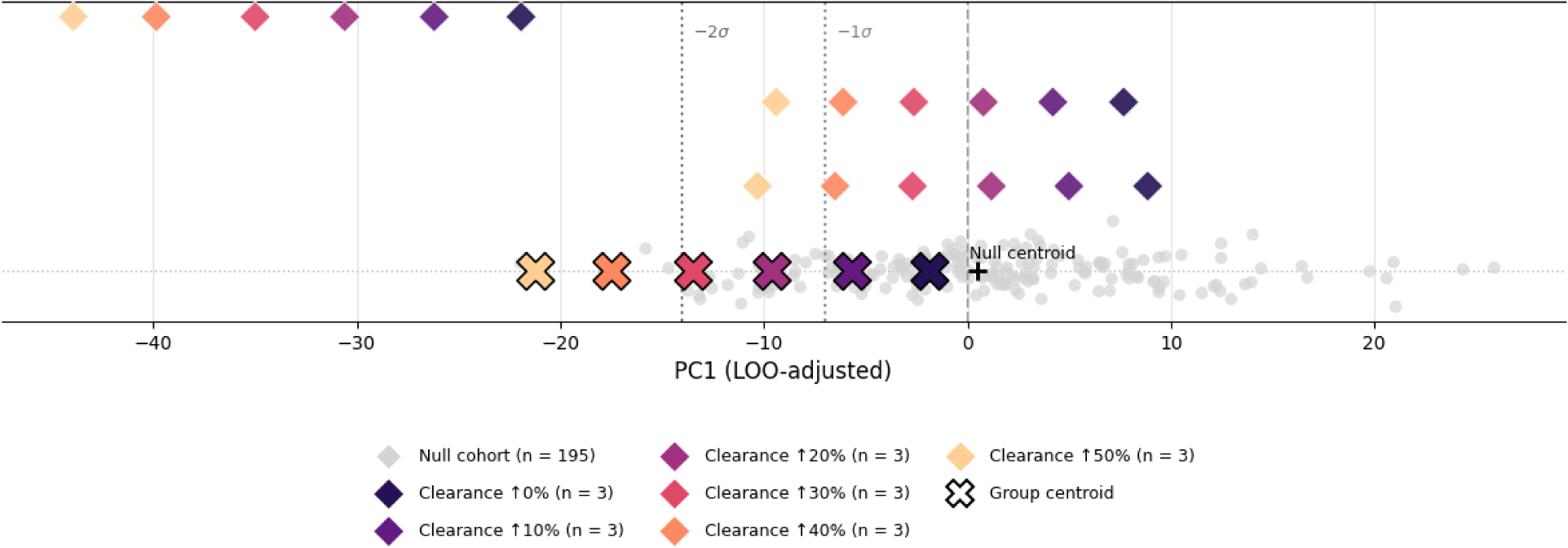
Semi-synthetic patient validation in PC1_adj_ space. Scatter of the n=130 null cohort with sufficient post-peak data to generate predictions in the simulated treatment time period (grey circles) overlaid with five permanent clearance-increase scenarios (10%, 20%, 30%, 40%, 50%; coloured diamonds, n=3 patients each). Group centroids of the clearance-increase scenarios (×) shift progressively leftwards in PC1_adj_ as treatment effect increases; the framework therefore identifies the clearance-mediated signal on the first principal component. Reference lines at −1σ and −2σ on PC1_adj_ are shown for context. Per-patient trajectories are provided in Supplementary Figure 9.

### Individual counterfactual residuals detect regenerative-therapy efficacy signals at smaller effect sizes and resolve them across dose levels

To explore the value of our framework for detecting treatment efficacy signals in comparison to current clinical trial approaches, we simulated a five-cohort dose-escalation trial (n=3 per cohort) under a regenerative-therapy mechanism. We compared the minimum detectable effect (MDE) of our framework to a published exemplar clinical trial protocol analysis which relies on interpreting biomarkers without deconvolution.(15) Across 2,000 resampled bootstrap trials per effect size, the framework reduced the MDE for the 80% power threshold from 67.5% (top-cohort clearance increase, planned analysis) to 24.5% (framework), a 2.76× improvement (Fig.6).

**Figure 6.**
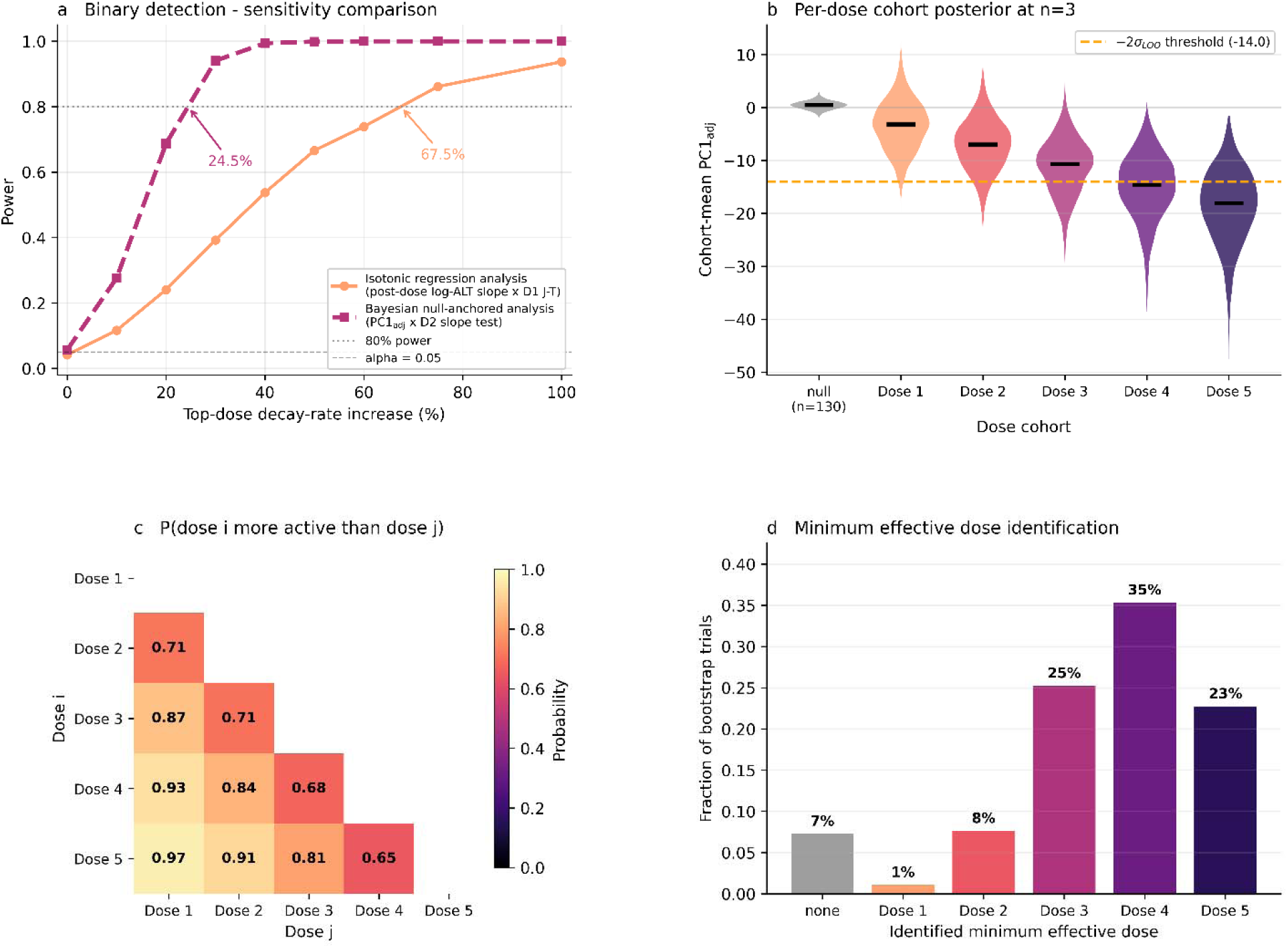
Statistical power and dose-finding support in phase 1 trial designs. (a) Minimum detectable effect (MDE): the framework reaches 80% power at a 24.5% increase in top-dose clearance; in contrast a published trial protocol analysis(15) reaches it only at 67.5%, a 2.76-fold reduction in MDE (2,000 bootstrap trials per effect size). (b) Per-dose bootstrap distributions of PC1_adj_ at the 80%-power top-dose effect size, sorted by assigned dose; the bootstrap-mean PC1_adj_ falls monotonically across dose levels. (c) Pairwise dose-ranking matrix: posterior probability that dose i exceeds dose j across 2,000 bootstrap trials. Under a monotonic dose-response (simulated), all cells converge above 0.5; under a plateau, cells comparing doses at and above the plateau would sit near 0.5 while comparisons against any below-plateau dose remain above the per-pair confidence threshold, identifying the inflection. (d) Minimum effective dose identification: histogram across bootstrap trials of the dose at which the per-dose posterior first crosses the detection threshold (i.e. the lowest dose crossing the −2σ_LOO_line in panel (b)). Full bootstrap procedure, test conventions and curve definitions in Supplementary Methods 5.

In each simulated trial, 15 patients were sampled and assigned across five dose cohorts (n = 3 each); a clearance-accelerating treatment effect of increasing magnitude was applied to their trajectories, and the remaining 115 patients were retained as an untreated dose-zero anchor. The historic controls serve as a valid dose-zero anchor under our framework because the within-patient counterfactual and LOO adjustment remove the patient-level variance (baseline severity, peak ALT, peak timing, and each patient’s own clearance rate) that would otherwise make a historic cohort incomparable to a trial cohort. In contrast, the exemplar in the published protocol analysis scores raw decay slopes, which retain that variance, so the same historic patients cannot validly be pooled into it. The power difference therefore reflects the framework’s removal of confounding variance, not a simple increase in sample size.

On these simulated trials we scored the planned analysis of the published exemplar trial (a Jonckheere–Terpstra ordered-alternatives test on the per-patient anchored log-ALT decay slope, for 15 treated patients) against the framework’s permutation slope test of PC1_adj_ on dose, which additionally uses the 115 historic-null patients as the dose-zero anchor (n = 130).(15) Per-dose PC1_adj_ posteriors (Fig.6b), pairwise dose-ranking matrix (Fig.6c) and bootstrap minimum-effective-dose histogram (Fig.6d) together provide a shape-resolved view that the protocol’s binary verdict cannot.

Smaller dose-escalation studies therefore become decisively informative, relevant for rare or severe acute injuries where patient accrual is slow. The same scoring function can also be applied to single-arm registries or compassionate-use case series anchored on a historic cohort.

Beyond pass/fail detection, the framework supports treatment/intervention dose-decision-making. The per-dose bootstrap distributions shift monotonically with assigned dose (Fig.6b), the pairwise ranking matrix quantifies per-comparison confidence (P>0.95 for well-separated doses; Fig.6c), and the minimum effective dose histogram identifies dose 4 as the most likely minimum effective dose to deliver a treatment effect (35% of trials; Fig.6d). This supports phase-3 entry at the lowest dose meeting the effect threshold rather than the top dose.

A consequence of fitting the EMG to each patient’s pre-intervention data and predicting across the full post-intervention trajectory is that the counterfactual residual is estimated from the whole post-intervention window rather than from a single follow-up observation. The EMG never sees the post-intervention data; it is fitted only on pre-intervention observations and used to generate a predictive trajectory against which post-intervention observations are scored.

### EMG decomposition identifies pharmacodynamic signatures consistent with pre-clinical studies

To establish the broader capabilities of our model for informing trial design using real-world evidence, we applied the EMG framework to four published case-series patients given fomepizole as an adjunct to N-acetylcysteine (NAC) for paracetamol poisoning, fitting all datapoints, rather than only post-intervention datapoints, due to the sparsity of pre-intervention data.(24) All four patients had peak ALT greater than 1,000U/L, the same severity threshold used to establish the null cohort (comparator-cohort sensitivity analysis in Supplementary Analysis 3).

NAC replenishes hepatic glutathione, which neutralises the toxic paracetamol metabolite N-acetyl-p-benzoquinone imine (NAPQI). NAC is most effective when given early, before glutathione stores are exhausted. Fomepizole has two potentially relevant mechanisms, established in pre-clinical studies, which have led to interest in a potential role as an adjunct treatment. It inhibits the production of NAPQI, and it also inhibits c-Jun N-terminal kinase (JNK) signalling, which amplifies mitochondrial injury after the initial NAPQI insult.(25) This JNK-inhibition remains active at later stages of injury, where the efficacy of NAC is known to be reduced.(26,27) The need for more effective therapies has led to fomepizole being used in 6.3% of paracetamol overdose cases in the USA, despite there being no clinical trial evidence for its use.(28)

This known mechanistic biology can be used to generate a prediction regarding the behaviour of EMG parameters in humans. If fomepizole truncates ongoing injury, then in a treated patient the fitted injury duration σ should be shorter, and the time of peak injury T should occur earlier than the personalised cohort prediction would suggest it should be based on the observed peak ALT. The clearance time constant λ should not move systematically: neither mechanism acts on biomarker clearance. Detection should be stage-dependent. Patients dosed during established hepatotoxicity, where NAC alone has limited capacity, should show the σ and T shift. The shift should be less apparent in patients dosed in the early injury window, provided NAC has been given early enough to limit NAPQI-mediated injury.(29)

For each fomepizole patient, the EMG was fitted with the NAC-only population priors. Each posterior parameter was then compared against a personalised cohort prediction derived from a log-scale regression on the subset of patients from the null cohort with known paracetamol ingestion times (n=96 NAC-treated controls; see Methods and Supplementary Methods 6).

The four patients (P1, P2, P3, P4) received their first fomepizole doses at 24, 31, 32 and 13 hours after hospital presentation, and had observed peak ALT of 3,325, 3,555, 3,812 and 1,260U/L respectively. By clinical staging, P1, P2 and P3 were first dosed during established hepatotoxicity; P4 was first dosed in the early injury window.

For P1, P2 and P3, dosed during established hepatotoxicity, the injury-duration narrowing is supported (Fig.7d). For P4, dosed in the early injury window, neither signature is present. λ showed no consistent directional shift across the four patients (Supplementary Figure 10). This stage-dependent pattern (a signal in the three patients treated during established injury, none in the patient treated when NAC is known to be effective) matches the mechanistic prediction; the across-patient single-parameter (σ and T) composites are tabulated in Supplementary Table 5, with prior sensitivity in Supplementary Analysis 4.

**Figure 7.**
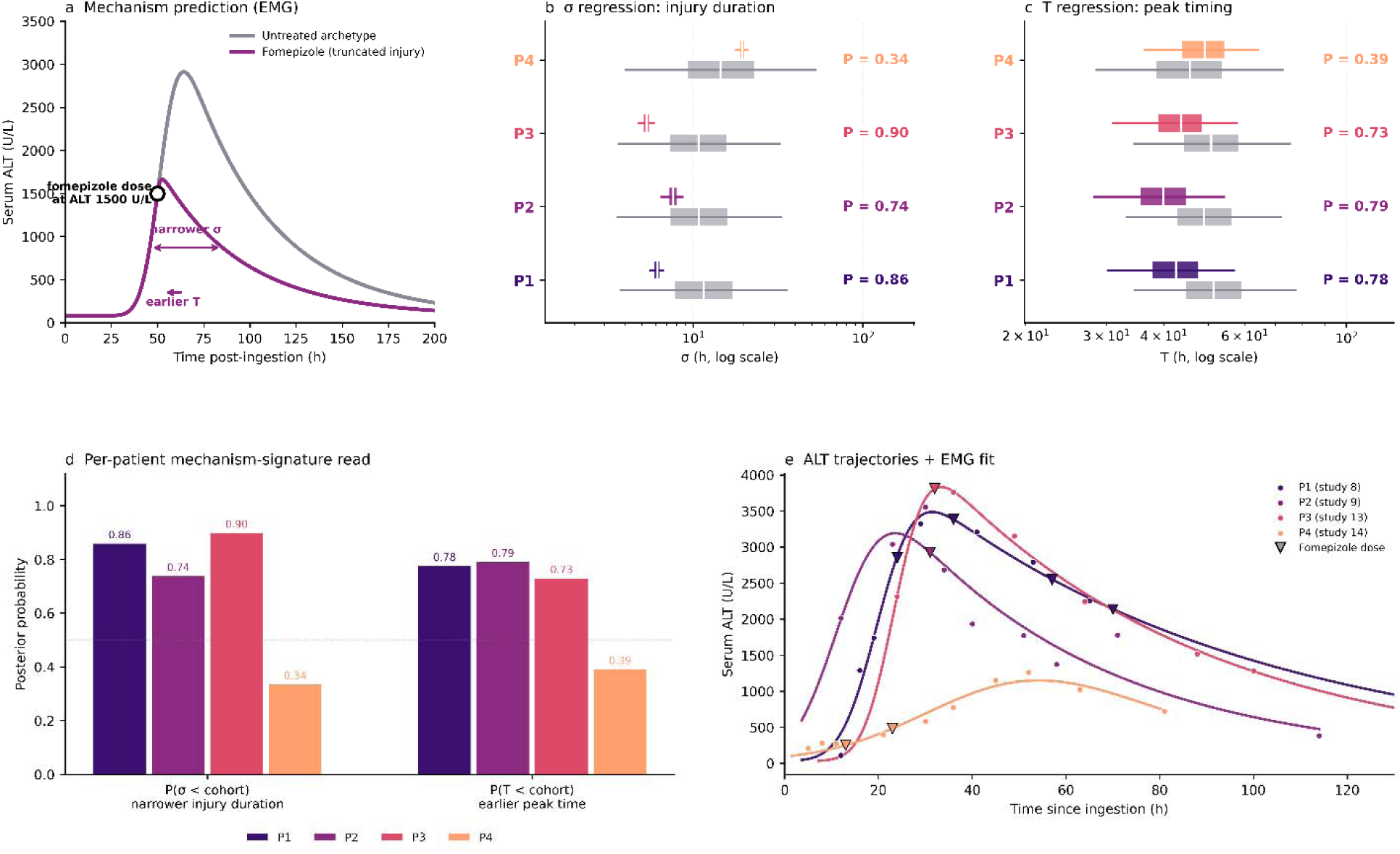
Fomepizole pharmacodynamic analysis. (a) Mechanism prediction: untreated cohort archetype (grey) compared with the predicted fomepizole curve (magenta) under CYP2E1- and JNK-mediated injury truncation, producing an earlier and lower peak with unchanged clearance. (b, c) Each fomepizole patient’s posterior σ (b) and T (c) against their personalised cohort predictive distribution (grey). (d) Per-patient signature: P(σ<cohort) and P(T<cohort) for P1-P4. Dotted line at P=0.5 marks the chance reference. (e) Observed ALT trajectories with EMG posterior-mean fits (the EMG is not fitted to the post-intervention data; the × marks the timing of fomepizole dose 1). Regression statistics and box-and-whisker conventions are in Supplementary Methods 6.

The framework reduces the sample size required to detect this effect prospectively. A confirmatory phase 2 trial of fomepizole, dosed under an established-hepatotoxicity trigger (peak ALT>1,000U/L) and powered to detect the σ-narrower signature at the per-patient rate observed in the treated case-series patients, would require n=16 patients at 80% power (Supplementary Analysis 5). The same study population in a naive ALT-change analysis would only be powered to detect an improbably high 86% treatment effect size at this number of participants, while a more realistic treatment assumption of a 42% treatment effect requires n=200. The framework’s sample-size advantage therefore extends beyond clearance modification, supporting its use as a unified inference approach across mechanism classes.

These data were analysed without retraining the model. The framework converts a published case series, from which raw ALT trajectories alone reveal only severe injury followed by recovery, into a per-patient mechanism signature that informs prospective trial design. As an uncontrolled case series subject to survivor-selection bias, this supports trial design rather than establishing fomepizole efficacy. However, the same retrospective framing extends across drug development: published case series and off-label registries currently treated as safety surveillance only have the potential to yield drug-class-specific pharmacodynamic signatures to inform later formal trial design.

## Discussion

The framework presented here addresses a common obstacle to efficacy inference in early-phase trials of acute organ injury therapies. It facilitates the interpretation of tissue-damage biomarker trajectories which are confounded by the temporal overlap of injury and clearance. Integration of Bayesian per-patient counterfactual scoring and functional PCA to accommodate sparse trajectories makes the use of historic controls as a dose-zero anchor feasible. The approach results in improved power to detect treatment effects, supports dose finding in small dose-escalation studies, and provides a route to detecting pharmacodynamic signatures to inform clinical trial design.

### Within-patient counterfactual scoring makes historic controls usable as a null population

In practical terms, the framework lets a phase 1 trialist with an uncontrolled dose-escalation cohort ask whether post-treatment trajectories deviate from each patient’s pre-treatment counterfactual at 2.76× smaller effect size than a standard slope-based analysis can detect. Historic controls re-enter as a dose-zero anchor rather than a level comparator on raw outcomes.(3,30)

Because the per⍰patient score, defined at the scoring step, already accounts for baseline severity, peak ALT, peak timing, and the patient’s posterior λ, the slope test becomes invariant to any drift in these quantities between the historical and trial cohorts.

### EMG deconvolution supports both clearance-modifying and injury-modifying inference

While the individual analytical components (Bayesian hierarchical modelling, EMG decomposition, functional PCA for sparse data, LOO regression adjustment) are established; this study is the first to integrate them into an end-to-end pipeline that distinguishes clearance-modifying from injury-modifying mechanisms. A standard change-in-biomarker score cannot separate the two.

For a clearance-modifying drug (e.g. a macrophage cell therapy that would be predicted to accelerate ALT decay; the MAIL trial setting), the PC1_adj_ pipeline applies.(15) For an injury-modifying drug (e.g. fomepizole, postulated to truncate hepatocyte injury), the λ-conditioned regression on the injury parameters applies.(25) A standard change-in-ALT score cannot, even in principle, distinguish the two mechanisms.

The fomepizole case series illustrates this: raw ALT trajectories reveal only severe injury and recovery, while the framework reads out per-patient mechanism signatures conditioned on individual covariates. Patients dosed in established hepatotoxicity (P1-P3) show signatures consistent with the predicted CYP2E1-and-JNK-inhibition mechanism; the patient dosed in early injury (P4) shows none, consistent with being treated during the stage when standard antidote therapy is effective. A stage-homogeneous prospective trial (n=16; Supplementary Analysis 5) could yield a primary readout that excludes the null.

### Per-patient posteriors provide continuous decision support across dose levels

In acute organ injury trials, each patient is dosed at a different point along an evolving disease trajectory; the framework accommodates this by comparing each observed trajectory against an individualised counterfactual fitted from pre-treatment data.

The comparator published protocol analysis we used will return a single binary monotonicity verdict across treated cohorts.(15) The framework reported here instead produces per-patient and per-dose posteriors that can support continuous dose-decision-making. This framework not only satisfies the dose-finding paradigm endorsed by published regulatory guidance, but can also be implemented here for indications where biomarker-based efficacy inference has previously been intractable.(1,31)

### Sampling flexibility supports trial-protocol deviations and real-world-evidence applications

Real-world ALT measurement is driven by clinical concern rather than protocol, so trajectories are sparse and irregularly sampled. PACE characterises each patient’s residual trajectory from whatever observations exist, retaining patients whose sampling deviates from protocol and opening a route to real-world-evidence analyses of pre-existing acute-injury cohorts.

### The architecture generalises conceptually to other acute tissue injuries

The statistical machinery (PACE-fPCA, LOO regression adjustment, permutation slope test with historic anchor) is indication-agnostic; the EMG deconvolution may require modification. For example, cardiac troponin potentially requires a multi-gaussian release model to accommodate the rise in troponin seen following percutaneous coronary interventions. However, the architecture offers a general template for efficacy inference across acute organ injuries, for re-fitting where appropriate kinetic models and historical-control cohorts exist.

### Limitations

Our model infers each patient’s injury parameters from the ALT trajectory alone; the ingested dose, plasma paracetamol concentration, ingestion pattern and presentation time are not consistently recorded in routine-care data. While the pipeline propates their absence as wider posterior and counterfactual uncertainty rather than biasing the readout, explicit parameterisation is a natural extension to explore if future datasets capture them.

The ALT data were collected as part of routine clinical care; sampling frequency varies with clinical acuity, which may introduce informative observation bias. The counterfactual predictions assume the historical cohort is representative; propensity-based adjustment would strengthen future applications. The Gaussian injury assumption may not hold for all presentations, but individual misspecification is absorbed into the cross-validated residual distribution, so detection performance depends on population-level calibration across 195 patients (Supplementary Figures 11 and 12) rather than on a perfect mechanistic fit for every individual. The counterfactual requires at least one pre-treatment ALT observation to anchor each patient’s fit, with precision improving as the number and spacing of pre-treatment samples increase.

Informative observation bias is a particular concern: ALT sampling reflects clinical acuity, so trajectory shape and the parameters fitted from it may be informed by missingness patterns correlated with severity. Standard imputation approaches (MAR/MNAR sensitivity, inverse-probability-of-observation weighting) are applicable, though their precision is intrinsically limited at phase 1 sample sizes; a confirmatory trial using this framework should consider pre-specifying Missing At Random/Missing Not At Random sensitivity analyses.

The framework’s use of historic controls depends on the scoring function removing the leading patient-level confounders; it does not remove higher-order drift in residual trajectory shape. The LOO coefficients are themselves estimated from historic data, so trial-era drift in the PC1-vs-log(λ) relationship would mis-specify the adjustment. However, there is no biological reason to suspect this would be the case.

A second mechanistic limitation concerns the assumption of first-order clearance. At very high ALT concentrations the elimination capacity may saturate, producing zero-order clearance that the EMG mis-specifies. Our null cohort’s residual structure shows no systematic deviation justifying a saturable parameterisation, but a Michaelis-Menten or biexponential clearance term could replace λ for clinical conditions where supraphysiological concentrations are routine, with negligible change to the rest of the pipeline.

The deconvolution approach via the EMG is mechanistically specific to tissue-damage biomarkers released by injured tissue in proportion to damage. Cell-signalling molecules (e.g. TNFα, IFN-γ, IL-6) are produced by effector cells under regulatory control, follow different generative dynamics, and would likely require a different model if there was a desire to relate them to tissue injury, rather than a re-estimated clearance constant.(16)

The framework is demonstrated separately for clearance-acceleration (Fig.5) and for injury-truncation (Fig.7). The framework cannot be applied blind to a drug or injury with an unknown mechanism. Both the semi-synthetic validation (Fig.5) and the power analysis (Fig.6) impose the treatment effect as an acceleration of clearance (the signal PC1_adj_ is constructed to detect). They therefore characterise detection sensitivity under a mechanism-matched best-case effect, and do not test robustness to treatment effects of a different residual shape; the fomepizole application (an injury-truncation effect detected through σ and T) provides the complementary mechanism class.

The fomepizole analysis uses published case-series patient data and is reported as per-patient signatures organised by dose timing (Fig.7d), not as an across-patient composite. The case series is subject to survivor-selection bias; no claim of fomepizole efficacy, mortality reduction, or transplant-free survival benefit follows. The single-Gaussian EMG cannot explicitly represent injury truncated mid-rise, which may contribute marginally to P4’s absence of signature. A non-parametric peak-restricted sensitivity (Supplementary Analysis 6) confirms the regression-based severity adjustment. Validation with actual trial data will provide the definitive test of operating characteristics; use in patient management would require prospective external validation against clinical endpoints, as detecting smaller effects is a sensitivity property; not all such signals will translate into clinical benefit. Signals at the margin likely require independent clinical-benefit anchoring (e.g. patient-reported outcomes, transplant-free survival) before being taken forward.

### Conclusion

In summary, the framework turns routinely-collected biomarker data from uncontrolled early-phase trials into a substrate for quantitative efficacy inference. It lowers the MDE for a small-cohort dose-escalation by 2.76× relative to standard biomarker analysis, supports both clearance-modifying and injury-modifying mechanism inference, and accommodates sparse data. Adopted at trial-design stage, this would reduce dose-cohort sizes - and therefore time, cost and patient exposure - required to declare a phase 1 readout fit for taking a candidate therapeutic forward. Consequently, it has the potential to reduce the size of, and in some settings remove the need for, a separate phase 2 dose-finding trial.

## Methods

### Preclinical model

Fifty twelve-week-old male C57BL/6J mice (Charles River, UK) were studied; 45 received 350 mg/kg paracetamol intraperitoneally after a 14-h fast, following the published protocol of Starkey Lewis et al.(32), and five fasted-only controls were culled without paracetamol to confirm baseline ALT. One paracetamol-treated mouse found dead at 7 h was excluded, leaving 44 paracetamol-treated mice in the ALT AUC–necrosis analysis. Serial tail-vein ALT was collected at 8, 16, 24, 36 and 48 h post-dose; livers were harvested at scheduled or humane-endpoint cull and percentage centrilobular necrosis quantified from H&E sections using InForm trainable image segmentation software. Full details are in Supplementary Methods 1. Animal studies were approved by the University of Edinburgh Animal Welfare and Ethical Review Body (093-LFR-24).

### Human clinical data

Patients admitted to NHS Lothian hospitals (Royal Infirmary of Edinburgh, Western General, St John’s; 2008-2024) with paracetamol overdose and peak ALT>1,000U/L were identified from clinical records, and enriched by identification of paracetamol overdose admissions to the Scottish Liver Transplant Unit to ensure capture of the full severity spectrum. Inclusion criteria were discharge diagnosis of paracetamol-induced acute liver injury with peak observed ALT>1,000U/L (the established threshold for clinically significant hepatocellular injury(33,34)) and at least three serial ALT measurements. All available patients meeting these criteria (n=195) were included; patient flow in Supplementary Figure 7. All patients received N-acetylcysteine as standard of care.(35) Data transfer was approved by the NHS Lothian Caldicott Guardian (Ref. 24167) and the study by Edinburgh Medical School Research Ethics Committee (25-EMREC-070).

### Bayesian EMG model

The ALT trajectory was modelled as:

ALT(t)=baseline + A x σ x sqrt(pi/2) x exp(0.5 x (σ/λ)2 + (T - t)/λ) x erfc((λ x (T - t) + σ2) / (λ x σ x sqrt(2)))

where A is the peak ALT release rate, σ is the injury duration, T is the time of peak injury, λ is the clearance time constant, and baseline is the pre-injury ALT (fixed at 30U/L). This is the convolution of a Gaussian injury process N(T, σ²) with first-order clearance exp(-t/λ).(36) The model was fitted in PyMC (v5.21.1) using Hamiltonian Monte Carlo (NUTS sampler, 4 chains × 2,000 iterations, 1,000 tuning, target acceptance 0.99). Priors, sampler diagnostics, and inter-rater model checking are detailed in Supplementary Methods 2.

### Functional PCA of residuals

LogL residuals r(t)=logL(ALT_obs(t) / ALT_pred(t)) were projected by PACE-fPCA(23) at each observed timepoint, with time normalised to each patient’s ALT peak. Full PACE setup and bandwidth selection in Supplementary Methods 3.

### LOO regression adjustment

PC1 scores were regressed on log(λ) by leave-one-out (LOO) regression.

### Scoring function rationale

The framework’s per-patient score f(patient)=PC1_adj_ absorbs four patient-level confounders by construction (baseline severity, peak ALT, peak timing, and the population-mean clearance component), and the residual orthogonal to these covariates serves as the substrate for the slope test. Rationale and orthogonality proof in Supplementary Methods 4.

### Semi-synthetic patient validation

Three patient ALT trajectories not included in the n=195 null cohort were used as the substrate for a semi-synthetic per-patient detection test (Supplementary Figure 9). For each patient, the EMG model was fitted on their pre-dose data only to give the posterior-mean null prediction at every observed timepoint; five permanent clearance-rate increase scenarios (10%, 20%, 30%, 40%, 50%) were then applied to the patient’s post-dose trajectory by recomputing the simulated post-dose ALT under each accelerated clearance, preserving the patient’s actual pre-dose measurements and measurement noise. For each (patient × scenario) pair, the residual trajectory log₂(simulated / posterior-mean predicted) was projected into the PACE-fPCA space trained on the 195-patient null cohort, and the LOO-adjusted PC1 score PC1_adj_ = PC1 − (intercept + β · log λ) compared against the cohort-derived detection threshold of −2 σ_LOO (Supplementary Methods).

### Power analysis for phase 1 trial designs

A Monte-Carlo power simulation was run on a separate analysis pool of n=130 historic-cohort patients with sufficient post-treatment data. Each patient’s observed ALT trajectory was transformed by a clearance-rate-increase effect at every dose level in the sweep. Five proportional dose levels (10%, 20%, 30%, 40%, 50% of an overall effect-size multiplier m swept 0 to 2) were assumed. For each m, 2,000 bootstrap trials were assembled, each drawing 15 patients from the 130-patient pool without replacement and assigning 3 patients to each of the 5 dose groups. Each trial was scored by two analyses applied to the same 15 simulated treated patients: (i) the framework - the 15 treated patients plus the remaining 115 historic-cohort patients used as the dose-zero null anchor, scored by a permutation slope test of PC1_adj_ on dose (299 permutations per trial, one-sided α=0.05); and (ii) the MAIL-protocol-style comparator - the 15 treated patients only, no historic comparator, scored as described below. False-positive rates fell within the nominal 5% under the global null for both tests.

For implementation of the MAIL-protocol-style comparator, the MAIL Trial protocol pre-specifies isotonic regression of change in ALT from baseline to day 7 across dose groups.(15) Two technical substitutions were required to obtain a runnable power-calculation implementation of this protocol-style analysis on the simulated data: (i) because not every simulated patient has a day-7 ALT observation, “change from baseline to day 7” was approximated by the per-patient anchored log-ALT decay slope - the OLS slope of log(ALT) versus time fitted through the patient’s pre-treatment ALT at t=0 and all available post-treatment observations; (ii) isotonic regression does not, on its own, return a hypothesis-test p-value, so for power calculation we used the Jonckheere-Terpstra ordered-alternatives test across the five treated dose groups (one-sided α=0.05) - the standard non-parametric hypothesis test for a monotone dose-response, and the conventional hypothesis-test analogue of isotonic regression. Both substitutions are made to give the protocol-style analysis a power calculation on the available data; neither is specified by the MAIL protocol itself.

### Cohort-regression-adjusted analysis: fomepizole-treated patients

Serial ALT for the four fomepizole-treated patients was obtained from Link et al.(24) (cases 8, 9, 13, 14; 8-10 timepoints per patient). The EMG was fitted to each patient individually using the same NAC-only population priors as the main cohort, with convergence assessed by the same diagnostics. This model used null cohort residuals for a model fitting using all datapoints, rather than incorporating prediction uncertainty, as the aim was to interpret the effect of the therapeutic on all fitted parameters.

Three cohort-level OLS regressions were fitted on log-transformed posterior parameters across the subset of patients from the null cohort with known paracetamol ingestion times (n=96 NAC-treated controls). Per-patient posteriors were compared against the personalised cohort prediction per MCMC draw, propagating within-patient posterior, regression-coefficient, and residual variance. Regression statistics (R² values, residual SDs, multiplicative bands) and full derivation are in Supplementary Information.

### Statistics and reproducibility

Preclinical: n=49 mice (44 paracetamol-treated in the ALT AUC-necrosis regression; 5 fasted-only controls excluded). Allocation was sequential, not randomised as mice are co-housed; ALT quantification and necrosis scoring were blinded. Sample size was exploratory, not power-calculated. Human cohort: n=195 patients with 2,036 serial ALT observations (183 had sufficient post-peak data for FPCA; 130 had sufficient post-treatment data for the power-analysis null). All eligible patients were analysed with no post-hoc exclusions.

Power analyses used 2,000 bootstrap trials per effect size; permutation slope tests used 299 permutations per trial (minimum p=0.003). Two-sided tests were used unless directionality was pre-specified by biological hypothesis (one-sided slope test for treatment-effect detection).

Bayesian diagnostics (R□, ESS, divergences) are in Supplementary Table 6; parameter identifiability is shown in Supplementary Figure 13, and cross-validation consistency in Supplementary Table 4.

### Implementation

All analyses were performed in Python 3.10 using PyMC 5.21.1 with the No-U-Turn Sampler for posterior inference, ArviZ 0.17 for convergence diagnostics, NumPy 1.26 and pandas 2.1 for data manipulation, scikit-learn 1.4 for regression and cross-validation, and Matplotlib 3.8 for visualisation.

## Data availability

The anonymised clinical datasets that support the findings of this study are not publicly available owing to NHS Lothian Caldicott Guardian restrictions on the sharing of patient-identifiable or derived clinical data, but are conceivably available subject to data governance approvals and completion of appropriate data sharing agreements. Mouse ALT and histological measurements are provided in Supplementary Information.

## Code availability

Analysis code for the Bayesian EMG model, the PACE functional PCA implementation, the LOO regression adjustment, and the simulated-treatment validation and power analysis pipelines are archived by Zenodo under DOI 10.5281/zenodo.20918409.

## Acknowledgements

For the purpose of open access, the author has applied a Creative Commons Attribution (CC BY) licence to any author accepted manuscript version arising from this submission. We thank William Mungall, Michael Walls and Helen Bradshaw for technical assistance with the animal studies. Funding: UK Medical Research Council (MR/T044802/1); Chief Scientist Office, Scotland (PMAS/21/07).

## Author contributions

CH, AMK, JC, LJS: Conceptualization, Methodology. CH, JC, CP, AF, MC, JM, RA, KJS, MJL, LJS: Investigation. CH, AMK, LJS: Software, Validation, Formal analysis, Data curation, Visualization. CH, SJF, LJS: Resources. CH: Writing – original draft, Project administration. JWD, SJF, LJS: Supervision. CH, JWD, SJF: Funding acquisition. Writing – review and editing: CH, AMK, JC, CP, AF, MC, JM, RA, KJS, MJL, PSL, AR, CJW, JWD, SJF, LJS.

