## Supplementary for "Efficacy inference in early-phase non-controlled clinical trials via Bayesian biomarker deconvolution"

### Supplementary Figures

##### Supplementary Figure 1. Shoulder artefact schematic

The shoulder artefact arises because ongoing hepatocyte injury overlaps temporally with ALT clearance from the circulation. (a) Two hypothetical patients sharing the same clearance rate lambda and identical total Gaussian injury, differing only in injury duration sigma: the prolonged-injury patient (wide sigma) shows a pronounced shoulder, whereas the brief-injury patient does not. The shoulder is a feature of injury duration, not of clearance. (b) Consequence of fitting a naive single-exponential anchored at the observed peak (with lambda recovered from the late tail): even when lambda is correctly estimated from the late tail, the model systematically under-predicts ALT during the early post-peak window when the Gaussian release is still active. This is the counterfactual bias motivating the EMG formulation.

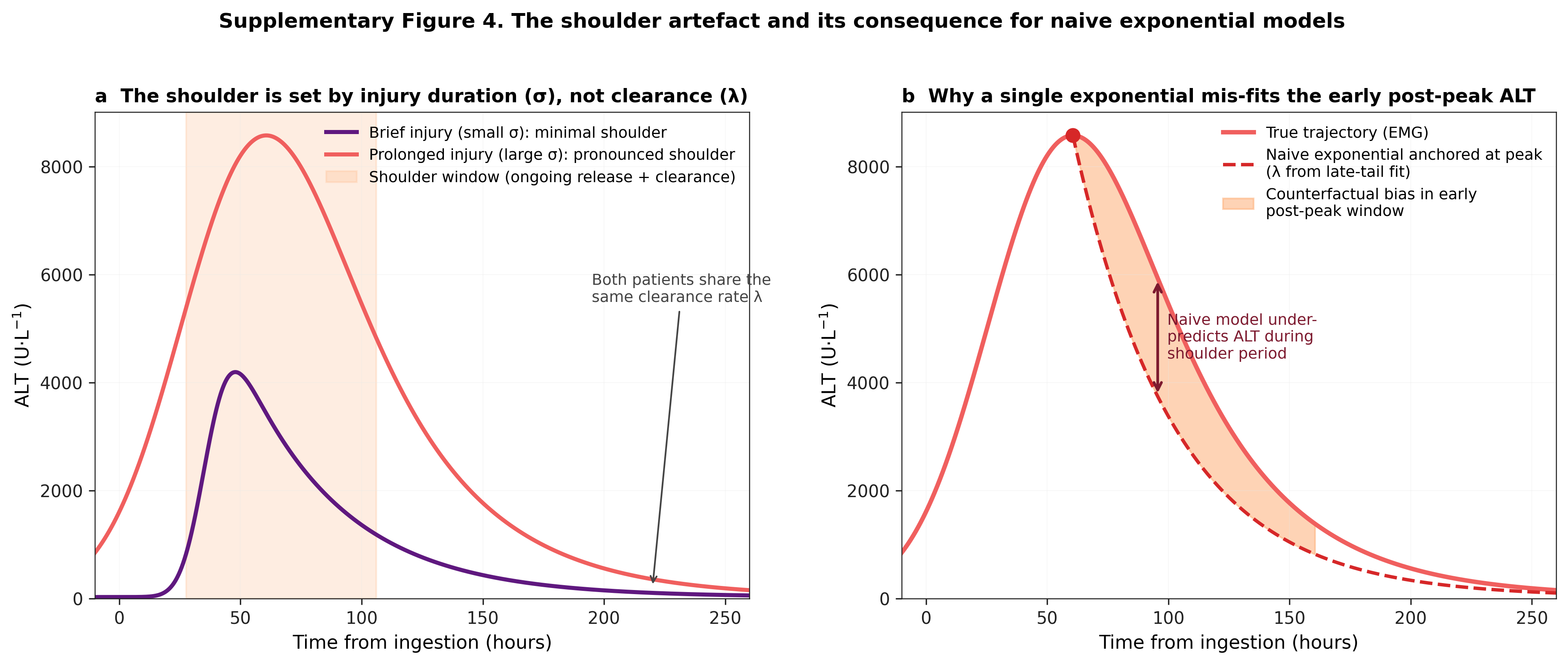

##### Supplementary Figure 2. Serial ALT for n = 44 mice treated with 350 mg/kg intraperitoneal acetaminophen.

Each line relates to one mouse, with a single marker at cull (final sample). Mice were sampled at baseline and at 8, 16, 24, 36 and 48 h until cull (n = 37); seven mice reached humane endpoint at unscheduled timepoints (11, 13, 14, 31 or 42 h, n = 7). Mice received 350 mg/kg intraperitoneal paracetamol. Serial tail-vein blood samples (40 microlitres) were collected at all timepoints until each mouse reached its scheduled or humane-endpoint cull point; a terminal sample was also obtained by cardiac puncture at cull, providing between 2 and 6 serial ALT measurements per mouse (including baseline). Mice culled at the earliest timepoint (8 hours) had two measurements (baseline and terminal), yielding the minimum calculable AUC.

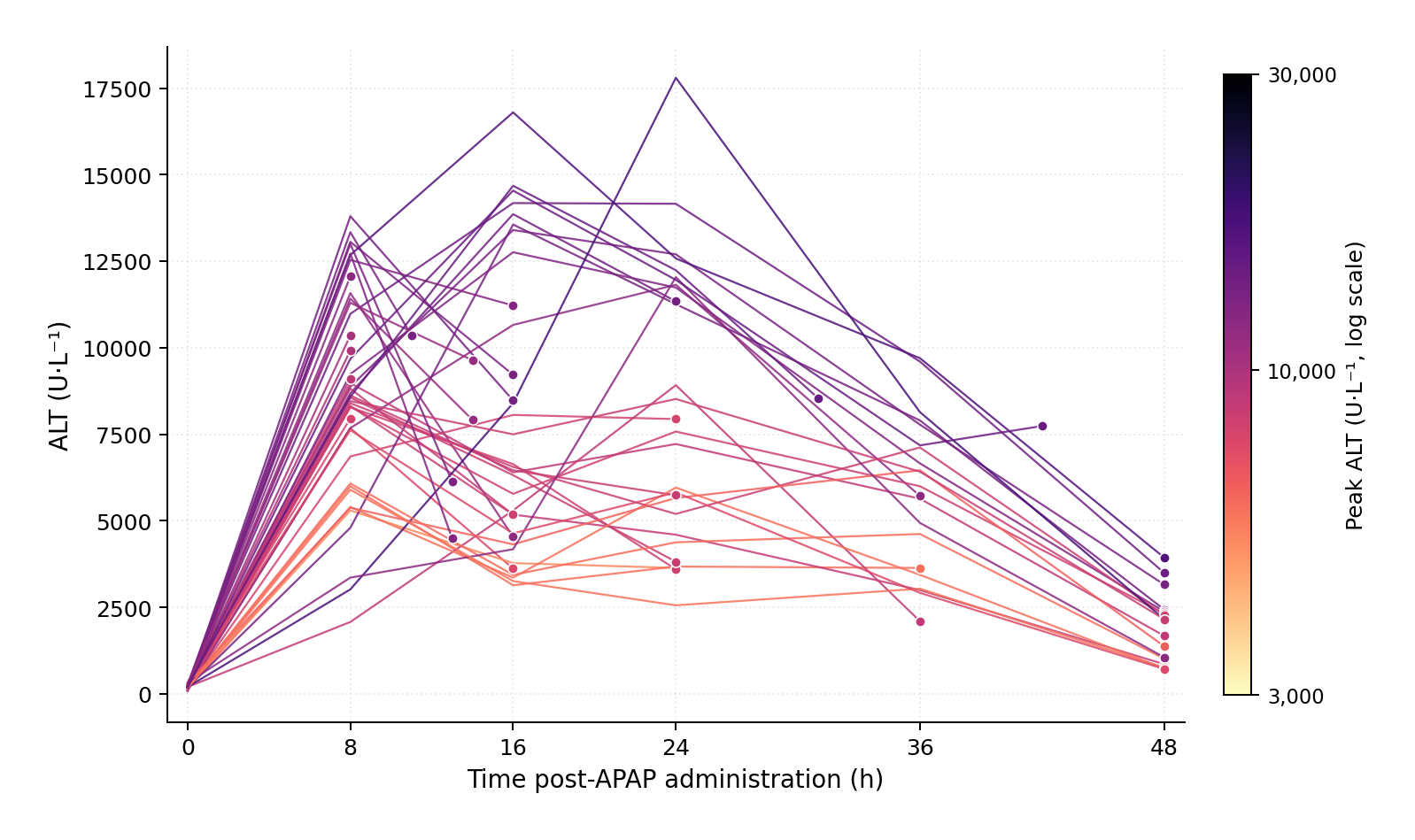

##### Supplementary Figure 3. Study flow

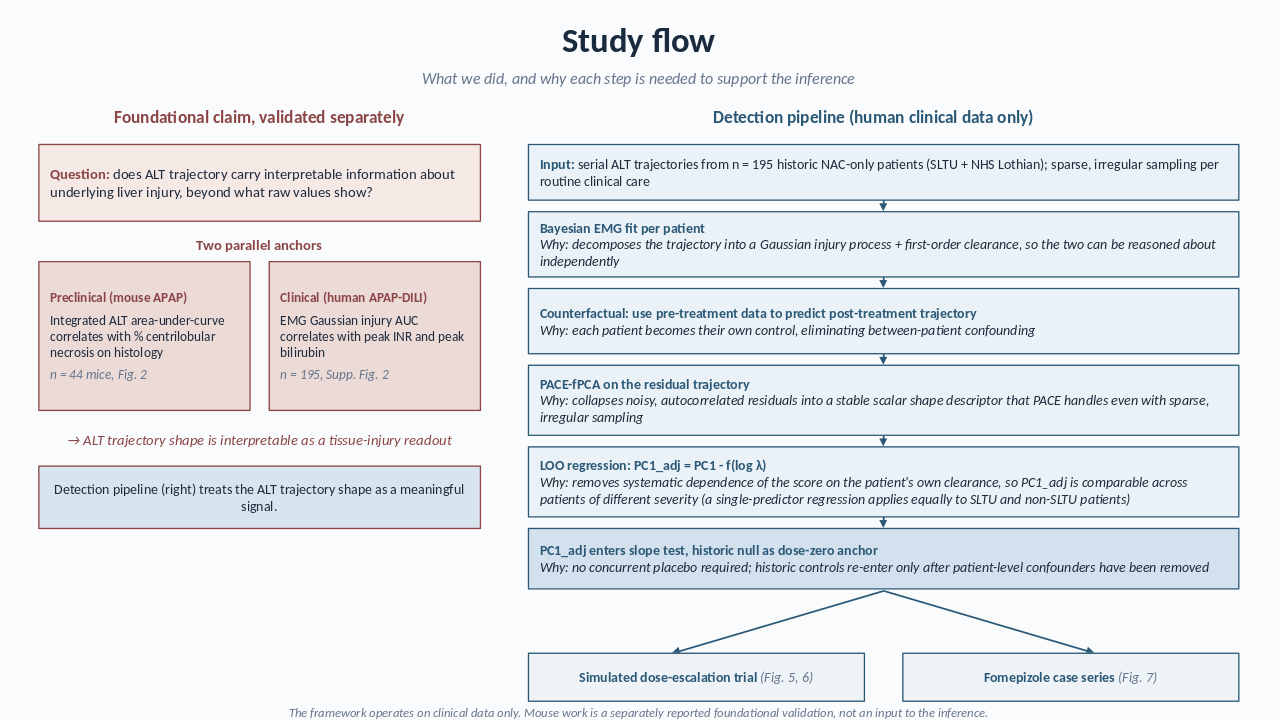

The framework operates on human clinical data only (right). The preclinical mouse work (left) and the human INR/bilirubin correlation are reported as a separate foundational validation that the ALT trajectory carries interpretable information about underlying liver injury; they do not feed into the detection pipeline. The EMG fit decomposes the trajectory so injury and clearance can be reasoned about independently; the counterfactual uses pre-treatment data to make each patient their own control; PACE-fPCA collapses noisy residuals into a stable shape descriptor; the LOO regression removes the dependence of the score on each patient's own kinetics; and then historic controls can be re-introduced as a dose-zero anchor for the slope test. The pipeline is applied to a simulated dose-escalation trial (Fig. 5, 6) and to a published fomepizole case series (Fig. 7).

The 195-patient historical-control cohort is used at several points: (i) fitting the per-patient EMG posteriors; (ii) training the PACE-fPCA eigenbasis; (iii) fitting the leave-one-out (LOO) regression that defines PC1_adj; (iv) providing the dose-zero anchor for the slope test in the power analysis; and (v) constructing the personalised cohort predictive distributions used in the fomepizole comparison. Two safeguards limit information leakage. First, the LOO construction ensures that no patient contributes to the regression coefficients used to compute its own adjusted score. Second, in every simulated trial the treated and dose-zero arms are disjoint subsets of the cohort, so no patient appears in both. The residual assumption is that the eigenbasis and the PC1–log λ relationship are stable between the historical-control cohort and a future trial cohort; trial-era drift in either would mis-specify the adjustment (see Limitations).

##### Supplementary Figure 4. Example automated tissue segmentation using InForm software, following segmentation algorithm training.

Following haematoxylin and eosin staining, necrosis was quantified as percentage necrotic area across a minimum of 10 digitally defined regions of interest per mouse, delineated using InForm tissue segmentation software (Akoya Biosciences). For each section, InForm defined subset regions excluding vessel lumina and air spaces; percentage necrotic area was quantified within these regions by automated classification of H&E-stained tissue. Scoring was performed by a blinded operator.

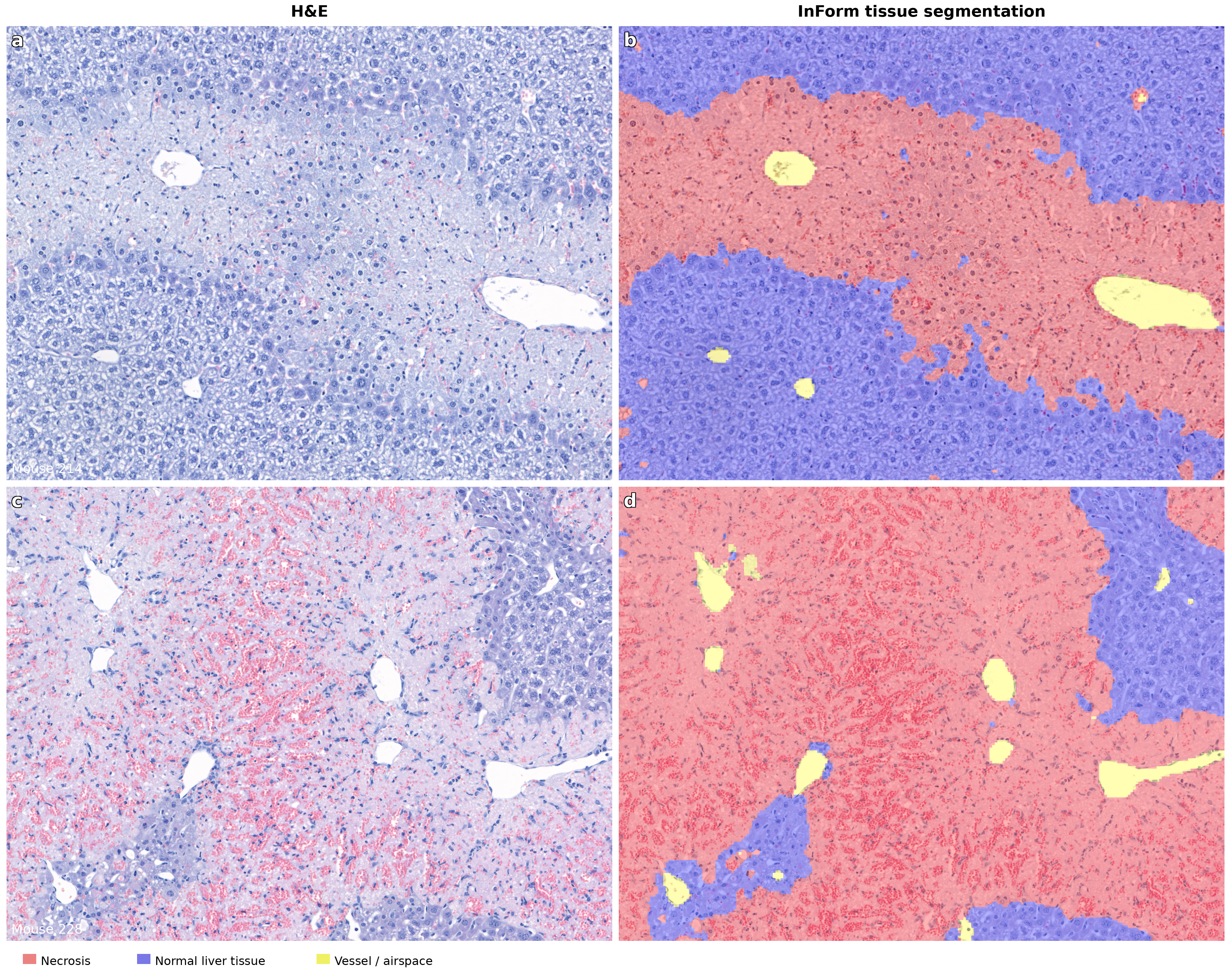

##### Supplementary Figure 5. Mouse log-log regression diagnostics

Residual diagnostics for the log-log linear regression model (necrosis = θ1 x time^{θ2} x ALT AUC). Six panels: (a) predicted vs observed necrosis, (b) Bland-Altman, (c) residuals vs fitted values, (d) Q-Q plot, (e) scale-location, (f) residual distribution. The Durbin-Watson statistic is not reported because these 44 mice represent independent cross-sectional measurements rather than a time series.

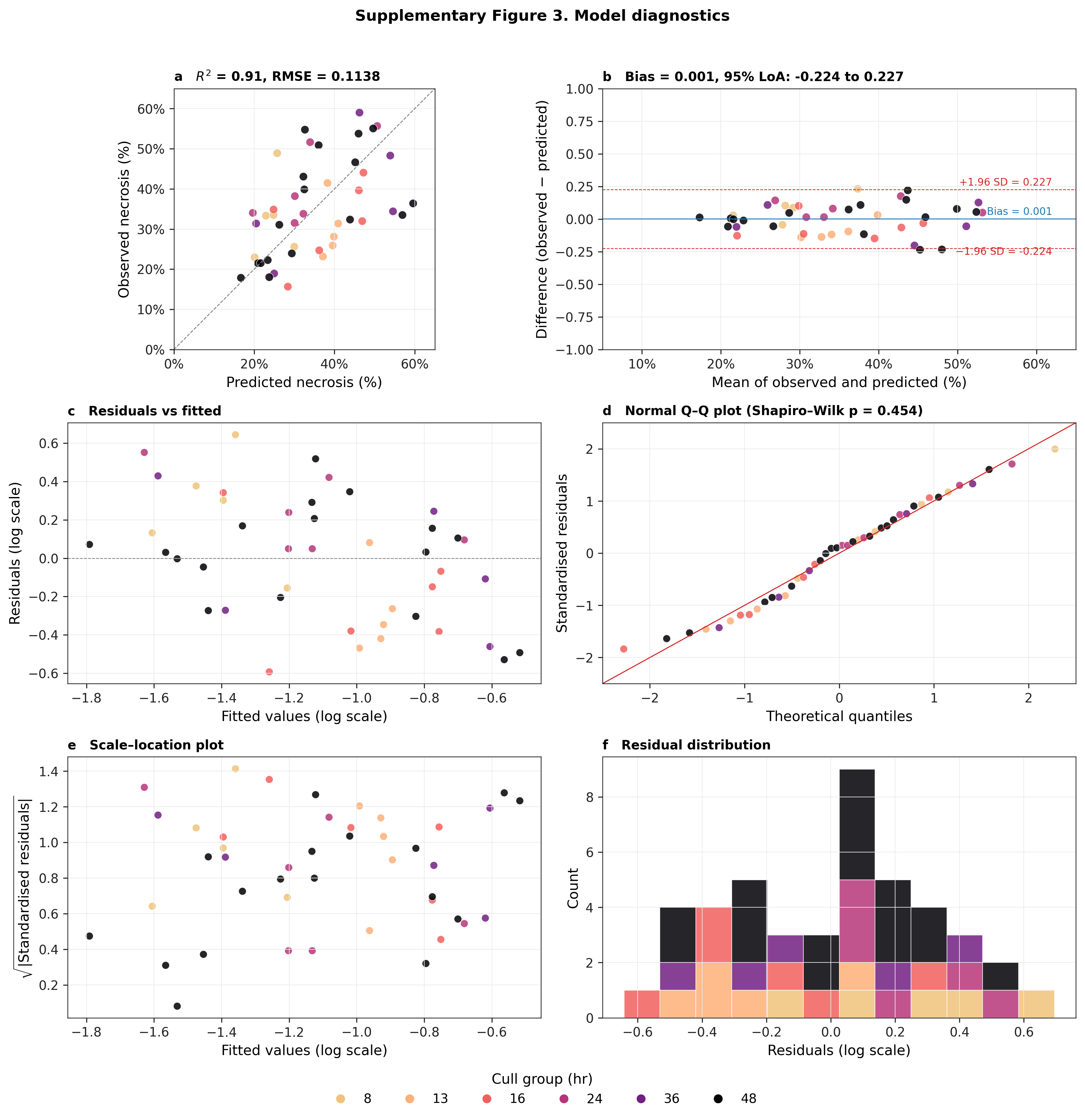

##### Supplementary Figure 6. Patient-level posterior means across the historical-control cohort (n = 195).

Histograms of per-patient posterior-mean estimates for the four inferred EMG shape parameters together with the inferred ingestion time (shown separately for the SLTU and NHS Lothian sub-cohorts, which use different ingestion-time priors) and the absolute measurement-noise scale σ_meas. Baseline y₀ is fixed at 30 U·L⁻¹ and is not shown. Panel annotations give the cohort median and interquartile range. Priors: A ~ Exponential(λ = 1/500); σ ~ Gamma(μ = 14, σ = 7); T ~ Gamma(μ = 60, σ = 12); λ ~ TruncatedNormal(50, 12; lower = 24, upper = 86); σ_meas ~ TruncatedNormal(4.576, 4.576; lower = 0.1, upper = 704); ingestion time ~ TruncatedNormal(0, 12; lower = −168, upper = first sample time) [SLTU] and TruncatedNormal(−51.83, 19.71; upper = −4) h [NHS].

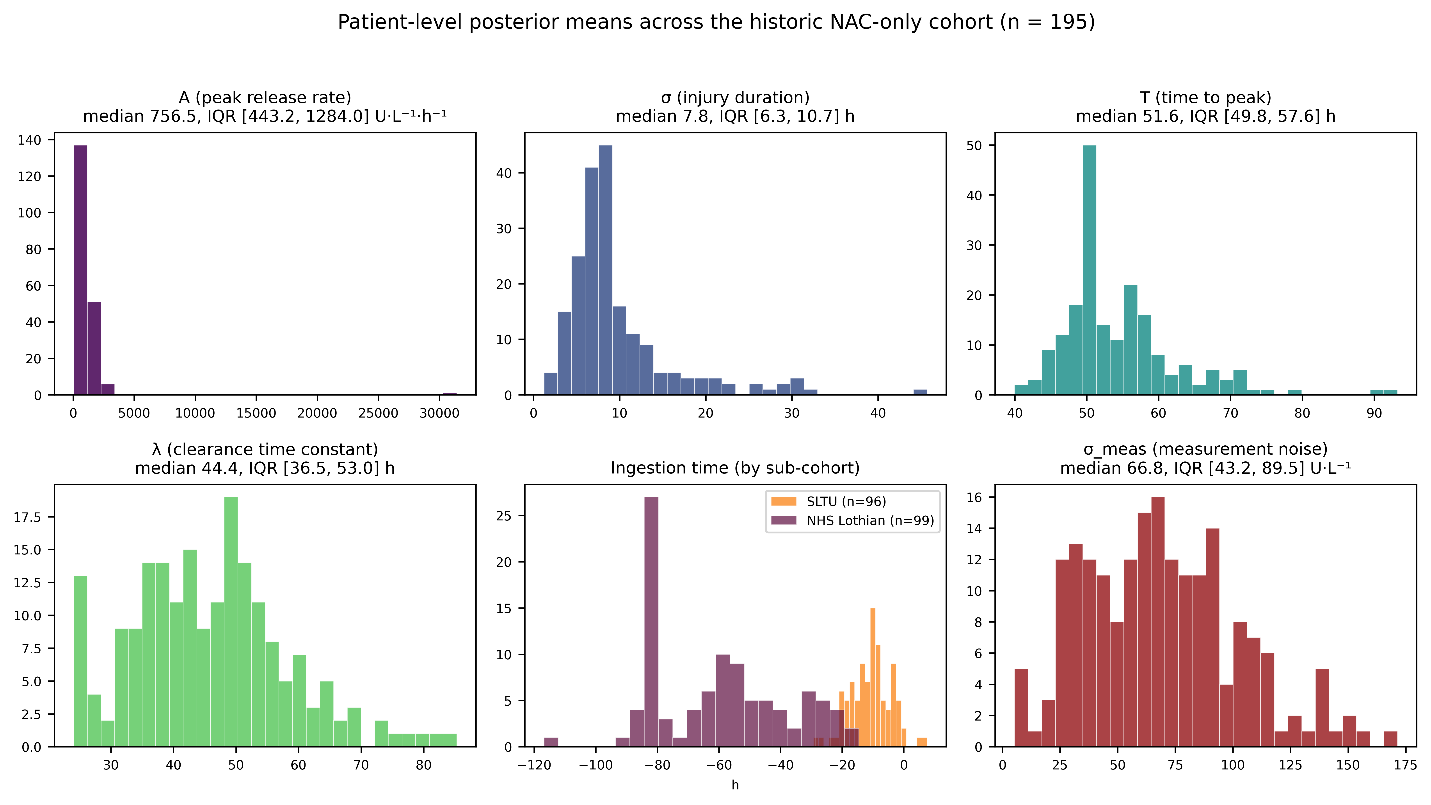

##### Supplementary Figure 7. Patient flow diagram

CONSORT-style identification and selection of the 195-patient APAP-DILI cohort. Patients were identified through two parallel strategies applied to NHS Lothian hospital admissions for paracetamol overdose: (i) ICD-10 = T39.1 cases identified from the hospital records system (n = 6,371), and (ii) identification from Scottish Liver Transplant Unit audit records (n = 100). Consistent inclusion criteria were applied to both source streams: at least three serial ALT measurements, peak ALT > 1,000 U/L during the index admission, and duplicate admissions for a single patient were removed. The resulting cohort of n = 195 patients comprises 96 SLTU patients (documented last-ingestion time, used in regressions that condition on a data-anchored ingestion convention) and 99 non-SLTU patients (ingestion time unknown, fitted with a prior). All patients received the antidote N-acetylcysteine as standard of care. The fomepizole case series (n = 4; Link et al., cases 8, 9, 13, 14) was analysed separately.

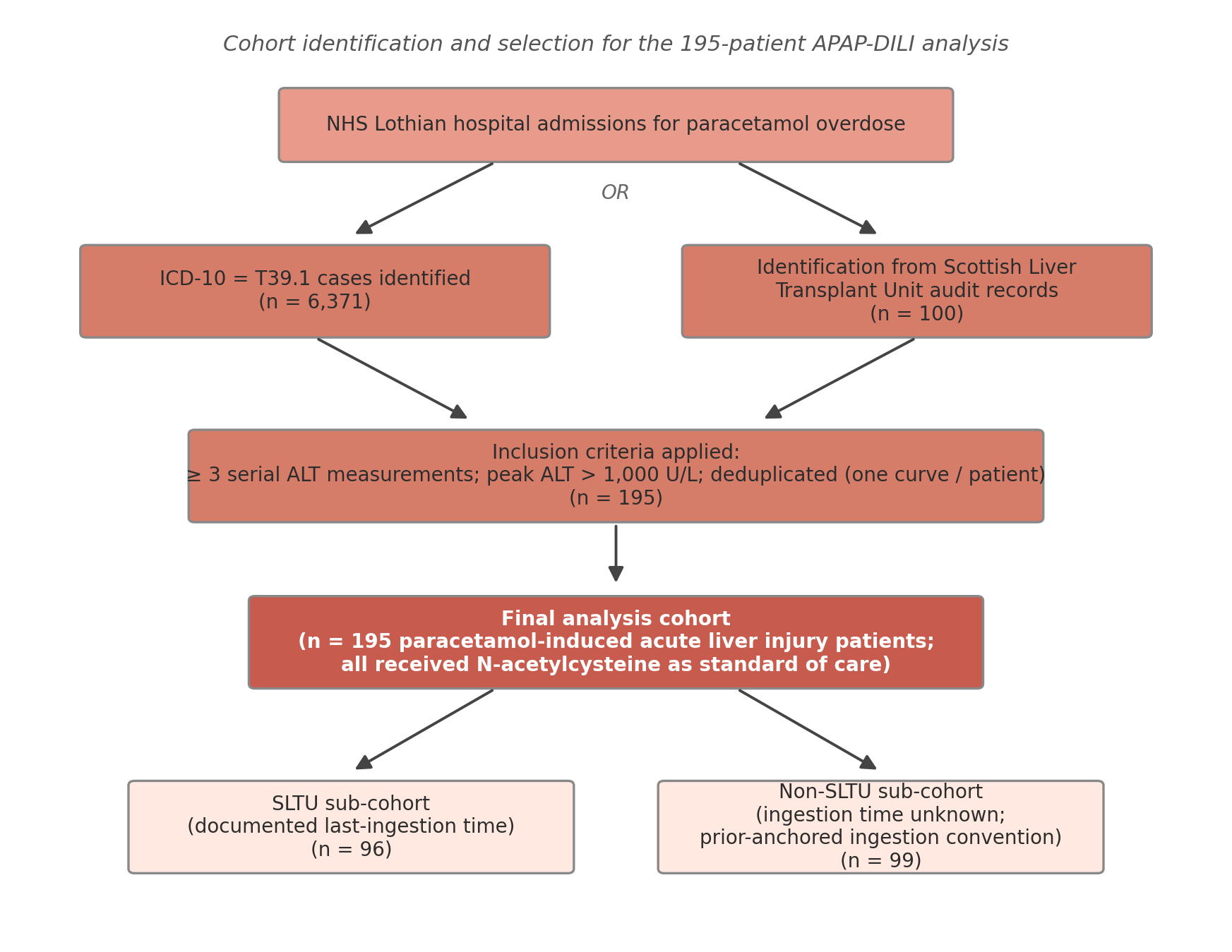

##### Supplementary Figure 8. Representative EMG patient fits

Full EMG posterior fits for six patients drawn from the cohort. Each panel shows the patient's observed ALT measurements (black markers, used in the fit) over the entire observed trajectory, the posterior-mean EMG curve from the full-data MCMC fit (coloured line), and the 95% credible band (shaded). Patient identifiers, fitted λ posterior means, and observation counts are shown in each panel header. y-axis on log scale.

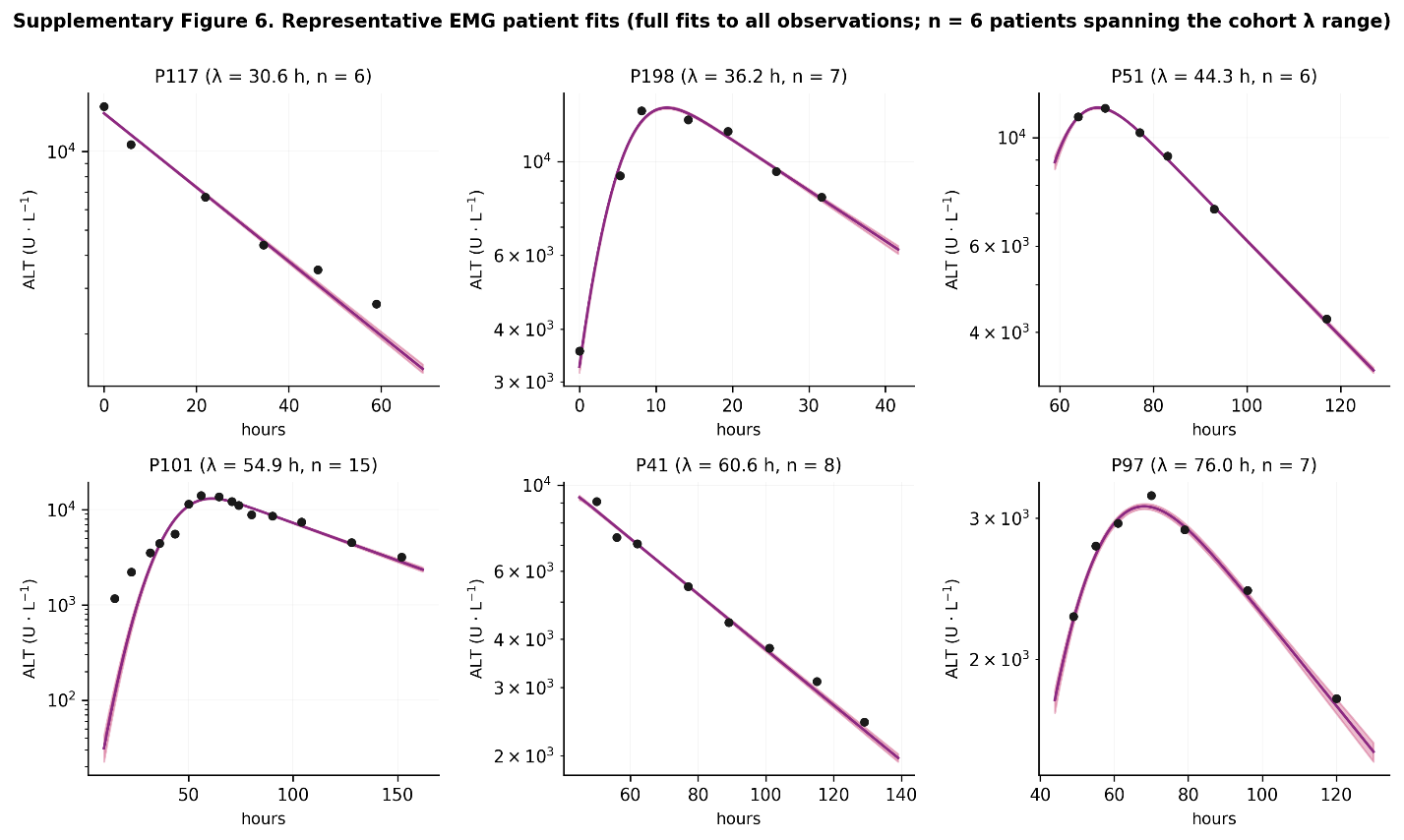

##### Supplementary Figure 9. Clearance-rate increase scenarios per real-trajectory simulated patient

Three real ALT trajectories were obtained from patients not included in the n=195 null cohort. For each patient, their null trajectory was their observed ALT data. Intervention scenarios simulated treatment effects once a 50% decrease from peak observed ALT was reached. The naïve decay rate was calculated from observed data at this timepoint, and then increased in 10% increments until a 50% increase. The simulated intervention timepoint for each patient sits at 63 h post-peak (P1), 32 h (P2) and 44 h (P3). Black markers are the patient's observed ALT measurements; the dashed grey line marks the intervention timepoint. The legend in the fourth panel identifies the scenarios. Clearance increases are recovered monotonically by the framework across all three patients spanning the cohort clearance range.

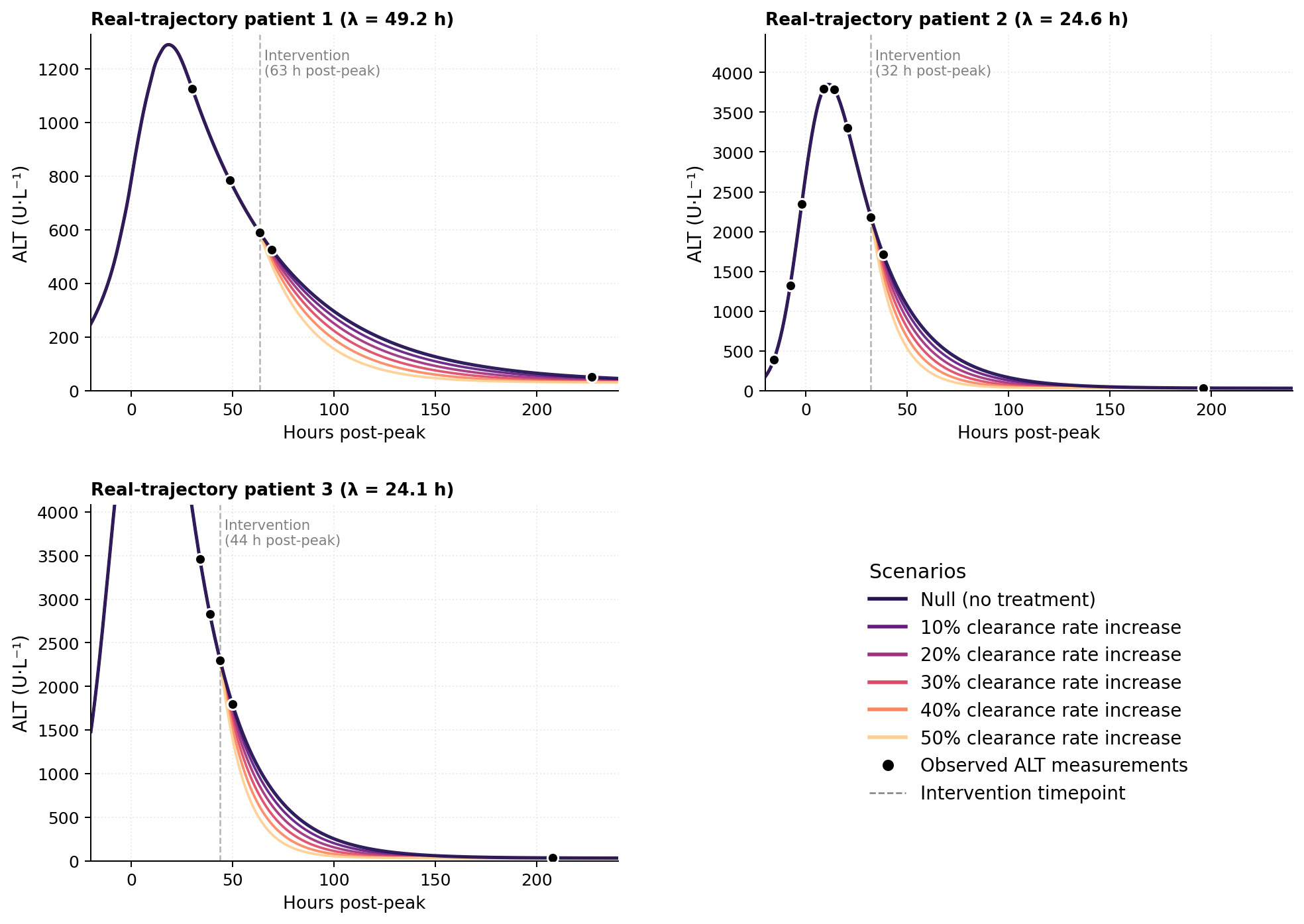

##### Supplementary Figure 10. λ-equivalence check for fomepizole

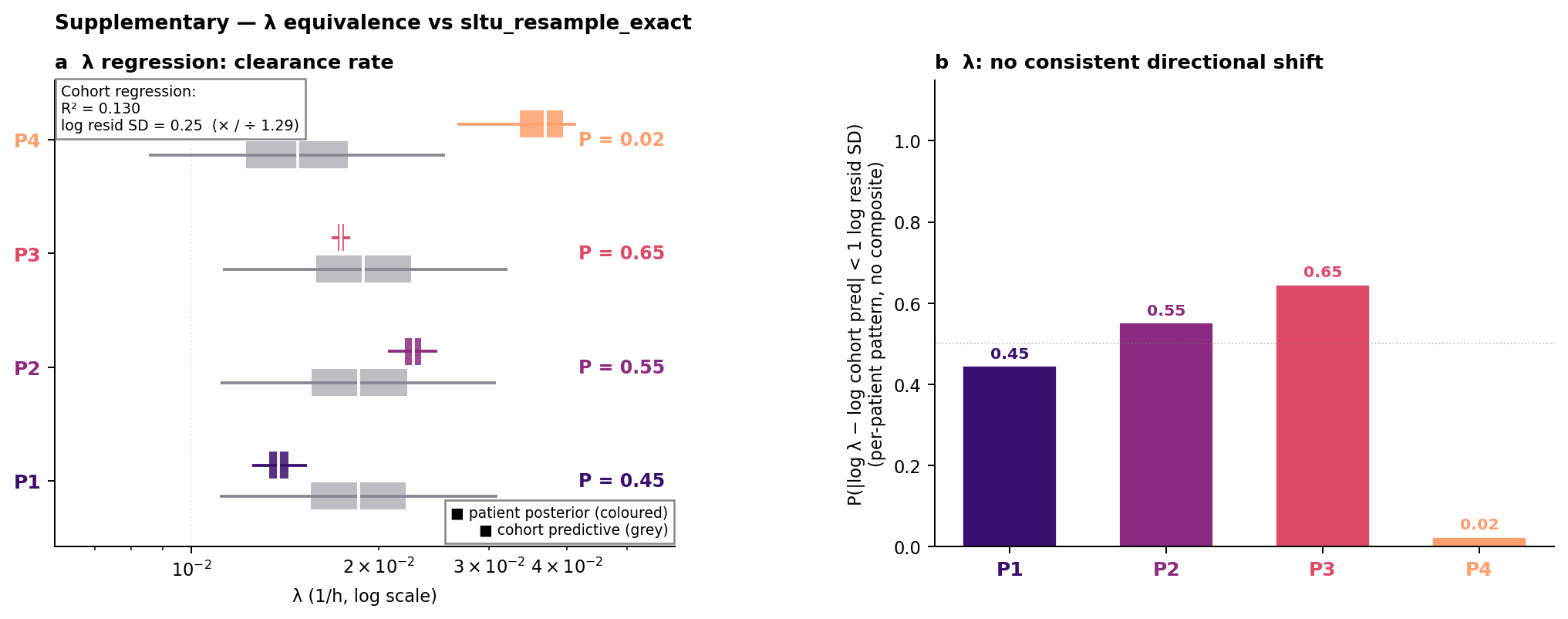

Per-patient check that fomepizole does not produce a consistent effect on ALT clearance. Panel (a) shows each fomepizole patient's posterior λ (coloured marker, with credible-interval whiskers) against the cohort predictive distribution (the SLTU cohort regression evaluated at this patient's own peak ALT, so each fomepizole patient is compared against the cohort expectation conditional on their individual severity; grey marker, with predictive-interval whiskers). The per-patient probability P that the patient's λ falls below the personalised cohort prediction is shown alongside. Panel (b) shows the per-patient probability that |log λ - log cohort prediction| < 1 log residual SD, an equivalence-style read. The pattern is multi-directional - P1 sits below the cohort prediction (slower clearance), P4 above (faster), and P2 and P3 within the equivalence band - consistent with no systematic clearance shift; we do not report a cohort-level Beta-Binomial equivalence composite because the per-patient direction is not consistent.

The framework reports the marginal benefit of fomepizole over the historical-control cohort regression, not the absolute injury-truncation effect of fomepizole. Because NAC is itself highly effective when started early after paracetamol ingestion, the marginal contribution of adding fomepizole is biologically expected to be small in early-treated patients and larger in patients treated once hepatotoxicity is established (where NAC's protective capacity is only partial, but the pre-clinically demonstrated mechanisms of fomepizole should conceivably continue to show benefit). The efficacy of NAC is typically considered to be negligible by 20-24hrs following ingestion of paracetamol.

P1, P2 and P3 were dosed 24-32 h post-presentation with peak ALT 3,325-3,812 U/L (established hepatotoxicity). Their posterior σ estimates fall in the 5–8 h range and posterior T in the 40–44 h range (Supplementary Table 4; Fig. 7b,c),with patient-level posterior probabilities supporting the σ-narrower and earlier-T signatures predicted by the CYP2E1 and JNK inhibition mechanism (SLTU comparator, errors-in-variables resample, exact mixture Beta-Binomial).

P4 was dosed at 13 h post-presentation with peak ALT 1,260 U/L (early injury, NAC-effective window). P4 has the widest posterior σ (19.5 h) and the latest posterior T (49 h) (Supplementary Table 4; Fig. 7b,c), with no σ-narrower or T-earlier signature attributable to fomepizole (P = 0.34 for the σ test, P = 0.39 for the T test). P4's λ sits below the cohort prediction (shorter decay time, faster clearance), in the opposite direction to P1 (whose λ sits above the cohort prediction); this is consistent with between-patient variance rather than a systematic effect (P = 0.02 for λ within the equivalence band). The absence of signature in P4 is consistent with NAC having provided protection against the toxic impact of ingestion, resulting in reduced opportunity for benefit from fomepizole.

In the absence of additional data to inform the decision, we would suggest that a prospective study should stratify on injury stage at first treatment and expect signature detectability only in the established-hepatotoxicity stratum. A secondary technical caveat: the single-Gaussian EMG cannot explicitly represent an injury cut off mid-rise, which may contribute marginally to broader fitted Gaussian parameters in early-treated cases.

##### Supplementary Figure 11. Residual diagnostics

Three-panel residual diagnostics computed across every observation in the 195-patient cohort (n = 2,036 individual log₁₀(observed/predicted) residuals; predicted ALT is each patient's posterior-mean EMG closed form evaluated at observation timepoints. (a) Residuals against posterior-mean predicted ALT (log x-axis), showing that relative residuals are most extreme at lower ALT values. (b) Residuals against time from each patient's posterior-mean peak (negative values are pre-peak observations, positive values are post-peak). Residuals shrink in the at-peak/early post-peak decay window before widening for late samples. (c) Residual distribution across all 2,036 observations. (Solid vertical line: cohort-mean residual (+0.094 log₁₀). Dashed vertical line: zero residual (perfect prediction reference). The offset between the two illustrates the positive bias driven by the long upper tail (panels a–b).

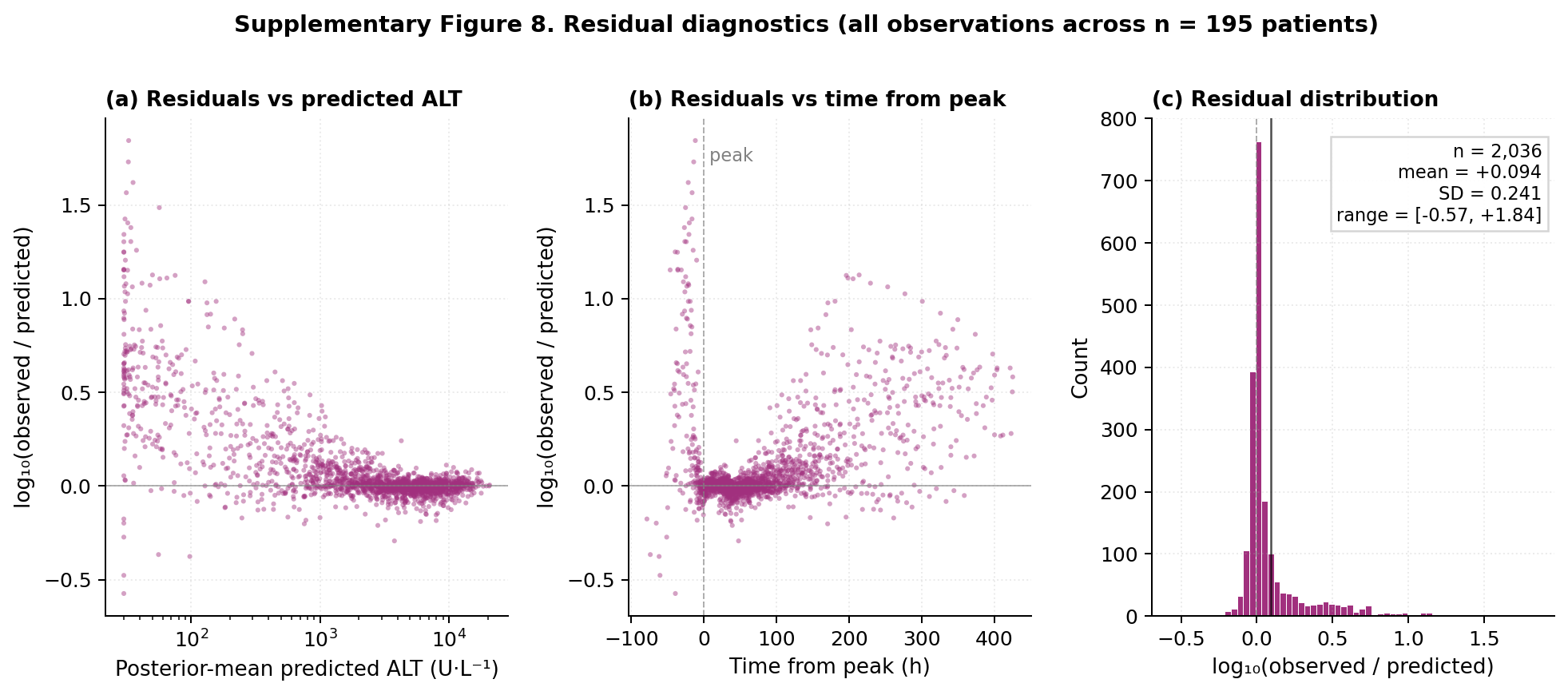

##### Supplementary Figure 12. Individual-level residual diagnostics

Per-patient log₁₀(observed / predicted) ALT residuals for nine patients sampled across the λ range, plotted against time from each patient's posterior-mean peak (peak time = ingestion time + peak time offset). Predicted ALT is the per-MCMC-sample mean EMG closed form from the full-data fit, evaluated at the patient's observed timepoints. Patient identifiers, fitted λ posterior means, and observation counts shown in each panel header; dashed vertical line marks t = 0 (peak time). Residuals fluctuate around 0 with magnitudes consistent with the measurement-noise component of the EMG model.

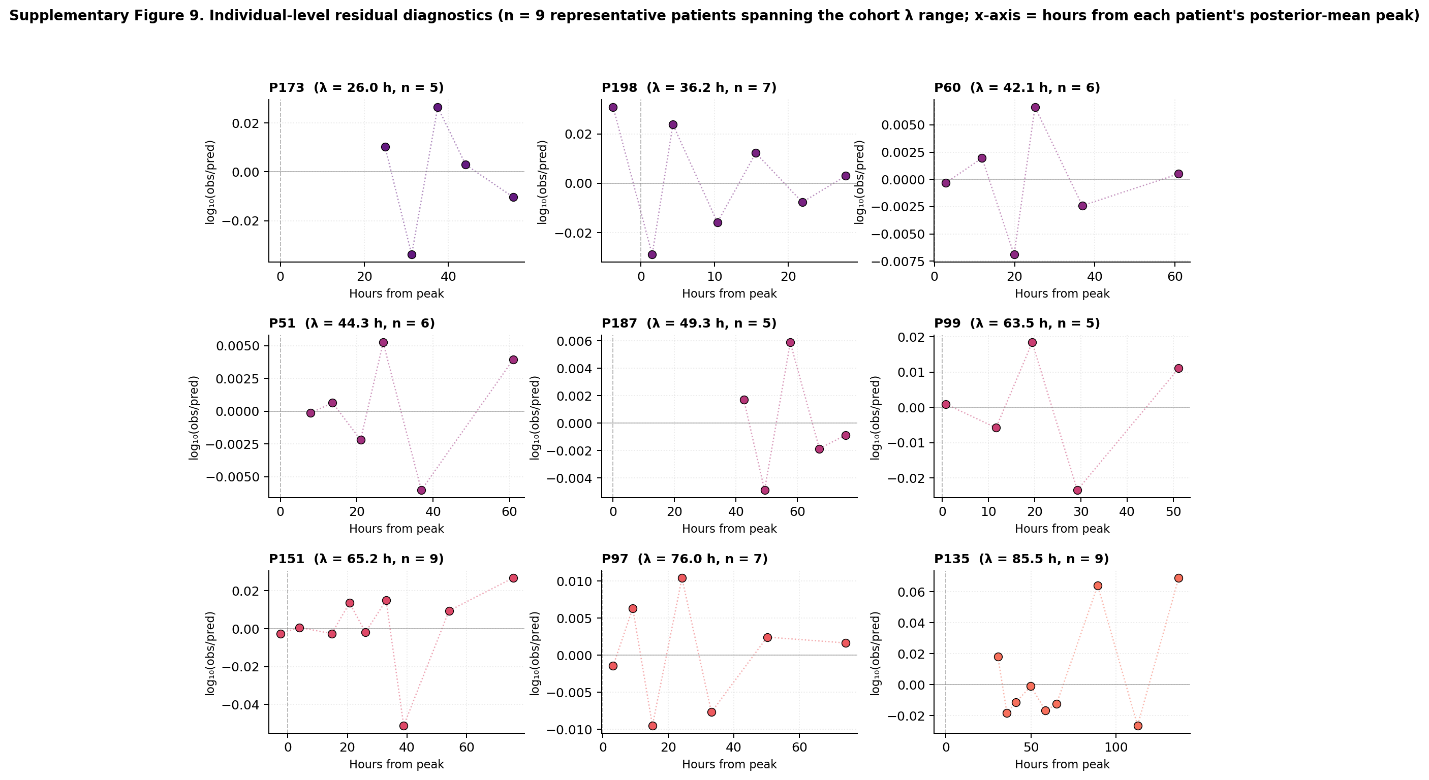

##### Supplementary Figure 13. Parameter identifiability diagnostics

Joint posterior MCMC density of A × σ for the same six representative patients shown in Supplementary Figure 8 (hexbin density; white circle marks the joint posterior median). The figure illustrates that the amplitude A and injury duration σ are partially identified per patient with the distribution reflecting both within-patient parameter correlation and the patient-specific data informativeness.

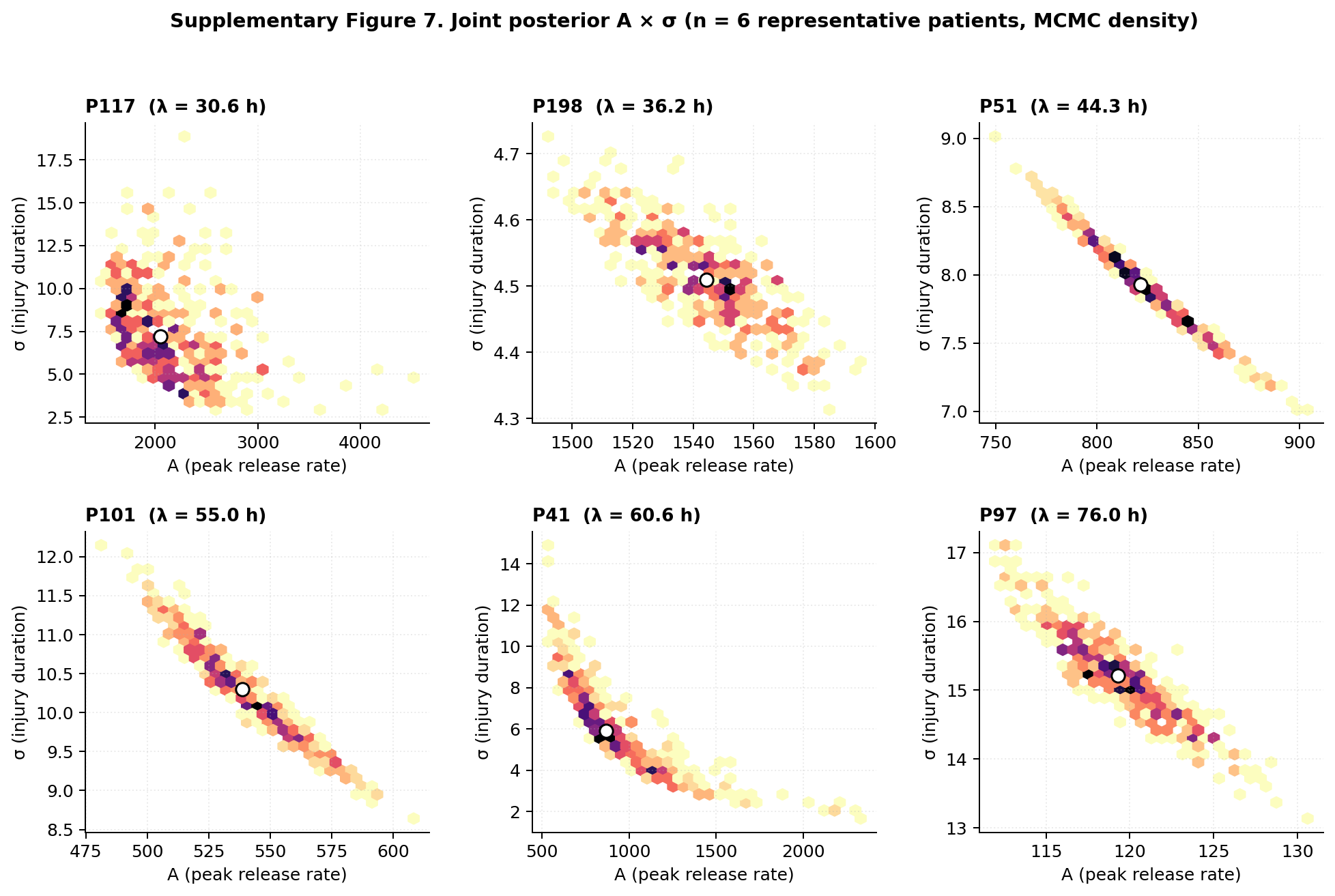

### Supplementary Analyses

#### Supplementary Analysis 1. Estimation of the ALT AUC mouse necrosis model

The power-decay structure of the ALT AUC-necrosis model was motivated by a two-step exploratory analysis: computing per-timepoint slopes (necrosis vs ALT AUC at each of six grouped cull timepoints), then fitting a power-decay curve to the change in gradient of those six slopes over time (Fig. 2a). The model parameters reported in the main text (θ_1_ and θ_2_) are estimated by ordinary least-squares regression on all 44 individual mouse data points.

The OLS model is fitted on the log-linearised form: log(necrosis / ALT AUC) = log(θ_1_) + θ_2_ · log(time) + ε. This constrains the ALT AUC exponent to 1 and estimates both parameters from every mouse. Both parameters were highly significant (θ_1_: p<0.001, θ_2_ p<0.001).

The OLS model constants were: θ_1_ = 4.074 × 10⁻⁵, θ_2_ = -0.914, R² = 0.91 (uncentred), RMSE = 0.114, bias < 0.01, 95% limits of agreement -0.22 to 0.23.

We also fitted an unconstrained model allowing the ALT AUC exponent to vary freely: log(necrosis) = α + β_1_ · log(time) + β_2_ · log(ALT AUC) + ε. The estimated ALT AUC exponent was 0.57 (SE 0.14), significantly different from 1.0 (F-test p=0.003), suggesting necrosis scales sub-linearly with cumulative ALT exposure. This three-parameter model improved prediction modestly (R² = 0.93, rather than 0.91) but at the cost of an additional fitted parameter without a clear rationale; we therefore report the constrained two-parameter OLS as the primary model.

#### Supplementary Analysis 2. Clinical relevance of the EMG-derived injury parameter

Does the EMG-derived Gaussian injury AUC capture clinically meaningful information about the severity of paracetamol-induced acute liver injury, beyond what the raw ALT trajectory shows?

We addressed this by correlating the Gaussian injury AUC with three established clinical severity readouts that could be supported by data available within the historical-control cohort: peak international normalised ratio (INR), peak bilirubin, and survival to discharge. INR captures loss of liver synthetic function; bilirubin captures excretory function loss; both develop downstream of (and on a longer timescale than) the ALT release window the Gaussian AUC characterises.

Across the historical-control cohort (n = 195), the gaussian injury AUC correlated positively with both peak INR (Spearman ρ = 0.255, p<0.001) and peak bilirubin (Spearman ρ = 0.258, p<0.001). Gaussian AUC did not separate survivors from non-survivors (Mann-Whitney U p = 0.73), which is not unexpected: mortality is a multiply confounded logistic outcome, while liver function is better reflected by INR and bilirubin. See Supplementary Figures 14, 15 and 16 for the per-readout scatter plots, survival comparison, and statistics.

The EMG-derived Gaussian injury AUC captures injury-severity information that is consistent with clinically validated severity markers in human paracetamol overdose.

##### Supplementary Figure 14. Gaussian injury AUC vs peak INR

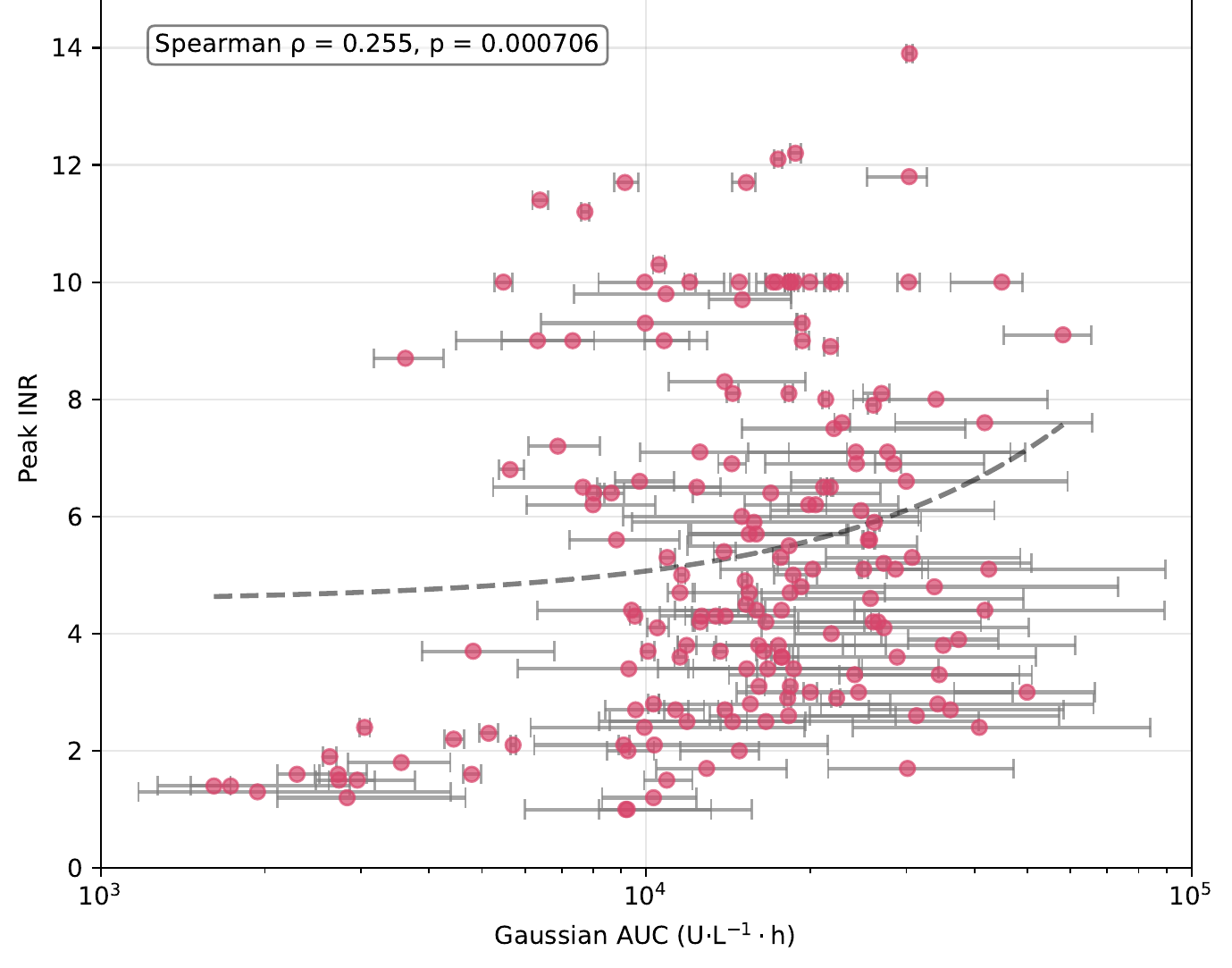

Scatter plot of EMG-derived Gaussian injury AUC against peak INR for the historical-control cohort (n = 195).

##### Supplementary Figure 15. Gaussian injury AUC vs peak bilirubin

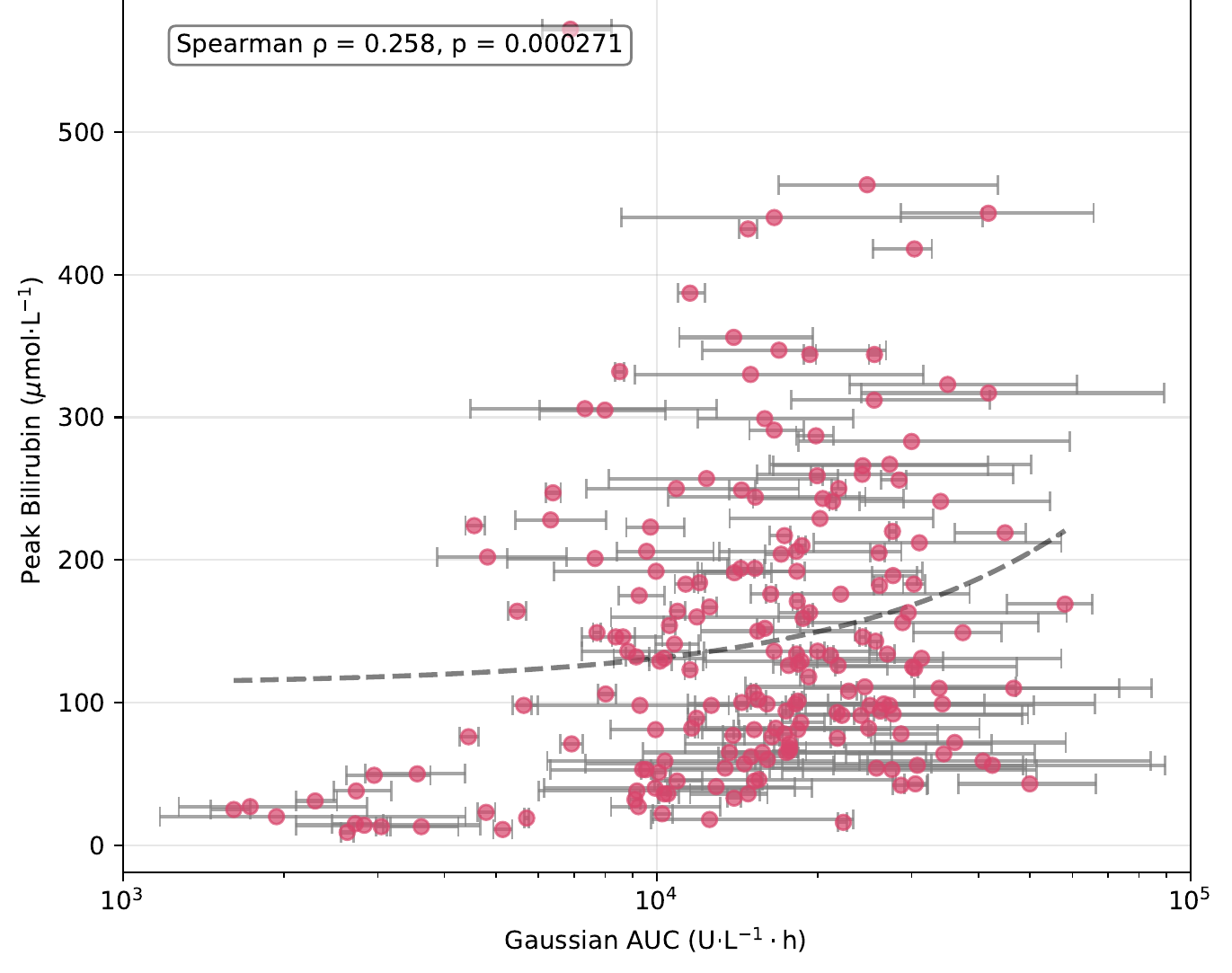

Scatter plot of EMG-derived Gaussian injury AUC against peak bilirubin for the historical-control cohort (n = 195).

##### Supplementary Figure 16. Gaussian injury AUC by survival to discharge

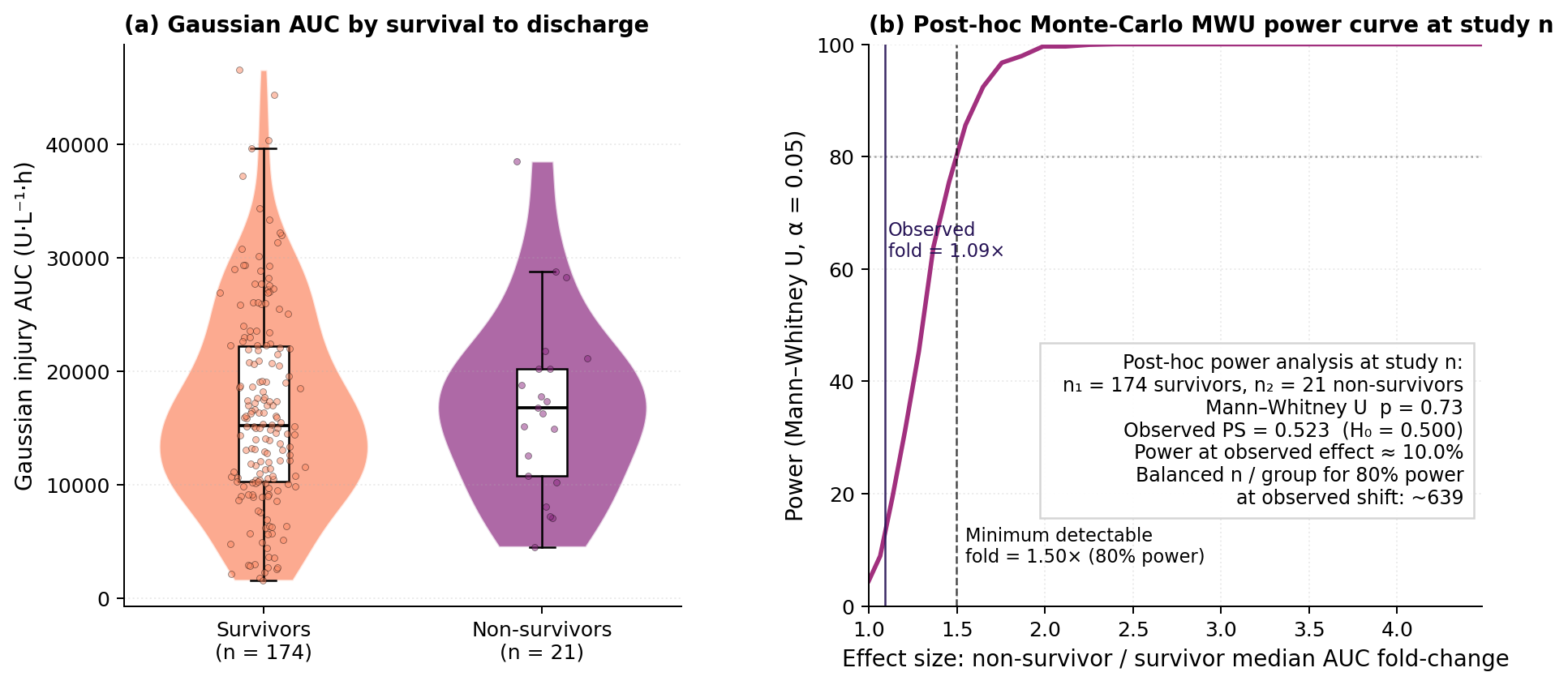

Survival to discharge comparison for the historical-control cohort (n = 195; n = 174 survivors, n = 21 non-survivors), with a post-hoc power analysis. Per-patient Gaussian injury AUC was computed from each patient's EMG posterior. (a): distribution of Gaussian injury AUC stratified by survival to discharge. (b): post-hoc power analysis at the study sample size. Mann-Whitney U on Gaussian AUC by survival to discharge gives p = 0.73; PS = 0.523 (H₀: 0.5). We cannot reject the null. Panel (b) translates the observed effect size into prospective-design terms: detecting a 1.10× fold-change at 80% power would require approximately 1238 patients across two arms. The combination of (a) and (b) shows that AUC-vs-survival does not separate in this cohort and the required study size to demonstrate separation is prohibitive.

#### Supplementary Analysis 3. Comparator cohort sensitivity

We re-ran the fomepizole comparison against three control cohorts: (i) SLTU only [headline, n = 96]; (ii) NHS only [n = 99]; (iii) pooled [n = 195]. The SLTU choice is retained because the data-anchored ingestion time matches the fomepizole patients.

#### Supplementary Analysis 4. Beta-Binomial prior sensitivity

The headline uses Beta(1, 1); we re-computed under Beta(0.5, 0.5) [Jeffreys] and Beta(2, 2) [weakly informative toward 0.5]. Composite means shift by ≤ 0.06 across priors for the σ and T marginals; CrI width contracts modestly under Beta(2, 2), as expected. The headline is robust to prior choice.

| Composite | Beta(0.5, 0.5) | Beta(1, 1) [headline] | Beta(2, 2) |
| --- | --- | --- | --- |
| σ < cohort | 0.66 [0.15, 0.99] | 0.64 [0.18, 0.97] | 0.60 [0.22, 0.92] |
| T < cohort | 0.63 [0.11, 0.99] | 0.62 [0.15, 0.97] | 0.59 [0.19, 0.92] |

#### Supplementary Analysis 5. Stage-homogeneous subset analysis: framework sensitivity is not compromised by low cohort R²

The R² for log σ on log peak ALT is 0.078 - high residual variance per patient, low for the headline regression. Here we aim to show that a small but clinically homogeneous subset can still yield a useful composite inference even at low R^2^.

For each fomepizole patient the framework reports a per-MCMC-draw posterior probability that the patient's σ is narrower than the cohort prediction at their observed peak ALT.

Under the assumption that a confirmatory established-hepatotoxicity stratum would be targeted at established injury, and therefore would have a per-patient probability of σ-narrowing equal to the P1-P3 pilot mean, the minimum prospective sample size at which the across-patient Beta-Binomial composite's 95% credible interval excludes 0.5 is n = 16 at 80% power (the smallest n such that power is at or above 0.80 for all subsequent n – supplementary figure 17). The corresponding T-earlier requirement is approximately n = 28. These projections assume a Beta(1, 1) prior, one-sided exact binomial inference against π₀ = 0.5, and a future per-patient success rate equal to the P1-P3 pilot mean.

##### Supplementary Figure 17. Sample-size and effect-size relationships for an EMG-informed clinical trial vs. a traditional trial of fomepizole.

Both panels show the relationship between the trial's pre-specified total sample size (x-axis, log scale 10–1000 patients in both panels) and the smallest treatment-related effect that the trial – designed at that fixed n – can reliably detect. Trial size is fixed in advance, and the y-value at each n shows the smallest true effect for which the trial would, in 80% of repetitions, reach its pre-specified conclusion of "an effect exists". The two designs cannot share a y-axis because they read out treatment effect on different scales (panel a: fractional reduction in peak-magnitude ALT; panel b: per-patient probability that the model resolves σ-narrowing), but the harmonised x-axis allows direct cross-comparison of patient-count economics at any given underlying mechanism strength. In both panels, yellow-green shading marks the (n, effect) regions in which the trial can detect the underlying effect; deep purple shading marks the regions in which it cannot. Vertical guide lines and matched annotation points at n = 16 (the framework's case-series anchor; rose orange) and n = 50, 100, 200, 500 (orange) allow read-across between panels.

The Bayesian decision rule rests on an integer success count. For a given *n*, the trial concludes "effect exists" when the observed positive-detection count *s* ≥ *s**(*n*), where *s**(*n*) is the smallest integer such that Beta(1 + *s**, 1 + *n* − *s**) excludes 0.5 from its lower 2.5% posterior tail. As *n* increases by 1, *s**(*n*) only changes at discrete thresholds; the minimum p_pilot at 80% power therefore steps abruptly each time *s** jumps. The envelope of the sawtooth is monotone decreasing toward 0.5 as *n* grows. Panel (a)'s curve is smooth by contrast because the t-test on continuous data has a continuous relationship between *n* and detectable effect size.

Panel (a) and panel (b) are not directly commensurable on the y-axis: a 50% Δ ALT reduction in panel (a) is a clinical-effect statement, while p_pilot = 0.83 in panel (b) is a model-resolvable detection-rate statement. The cross-comparison via shared n is therefore a statement about patient-count economics, not a statement about which design measures a larger clinical effect. The σ-narrowing endpoint is more powerful per patient at the same n because the model deconvolves a mechanism-specific feature (truncation of the injury timescale) that is diluted in a peak-magnitude 2-arm study.

Panel (a) - Naive randomised RCT. Two-arm 1:1 randomised, double-blind, placebo-controlled trial of fomepizole + NAC vs placebo + NAC, primary endpoint log Δ ALT (presentation → peak ALT), two-sample t-test against the null of no treatment difference. Design template based on a published trial.^1^ Y-axis: smallest detectable Δ ALT reduction (%) - the smallest fractional reduction in geometric-mean Δ ALT (treated vs control) for which the trial, designed at the x-axis n, achieves 80% power. Assumptions: (i) log Δ ALT is the per-patient endpoint; (ii) noise SD on the log scale is 1.39, the empirical SD of log Δ ALT in the 113-patient sub-cohort of our historic control cohort who were still on the rising limb of injury at first observed ALT - those who would have been eligible for randomisation under a presentation-trigger recruitment criterion; (iii) variance is assumed equal in treated and control arms; (iv) the trial recruits to its target and is analysed per protocol. Per-arm n is the total n divided by 2 (per 1:1 randomisation).

Panel (b) - Framework single-arm trial. Single-arm trial, all patients receive fomepizole. For each enrolled patient, the EMG model is fitted to their ALT trajectory and the per-patient σ-narrowing posterior probability P(σ_treated < σ_cohort) is computed against the historic 195-patient cohort posterior. A patient "counts" toward the trial endpoint when this posterior probability exceeds 0.5 (i.e., when the patient's data make it more probable than not that their treated σ is narrower than the cohort-predicted σ). Decision rule (Bayesian): the trial concludes that fomepizole has a treatment effect when the data give convincing evidence that the underlying per-patient detection rate is above chance - formally, when the 95% posterior credible interval on that rate excludes 0.5. Y-axis: smallest detectable per-patient detection rate p_pilot - the smallest underlying rate at which an EMG-fitted patient produces a σ-narrowing posterior probability exceeding 0.5, for which the trial, designed at the x-axis n, has at least 80% probability of reaching the decision rule. The y-axis is a model-resolvable detection rate, conditional on the typical data density and presentation timing matching the case-series patients in whom our main manuscript analysis suggests potential for treatment efficacy. The dashed yellow horizontal line marks the case-series-anchored pilot detection rate p_pilot = 0.83 (the mean of P1, P2, P3's σ-narrowing probabilities).

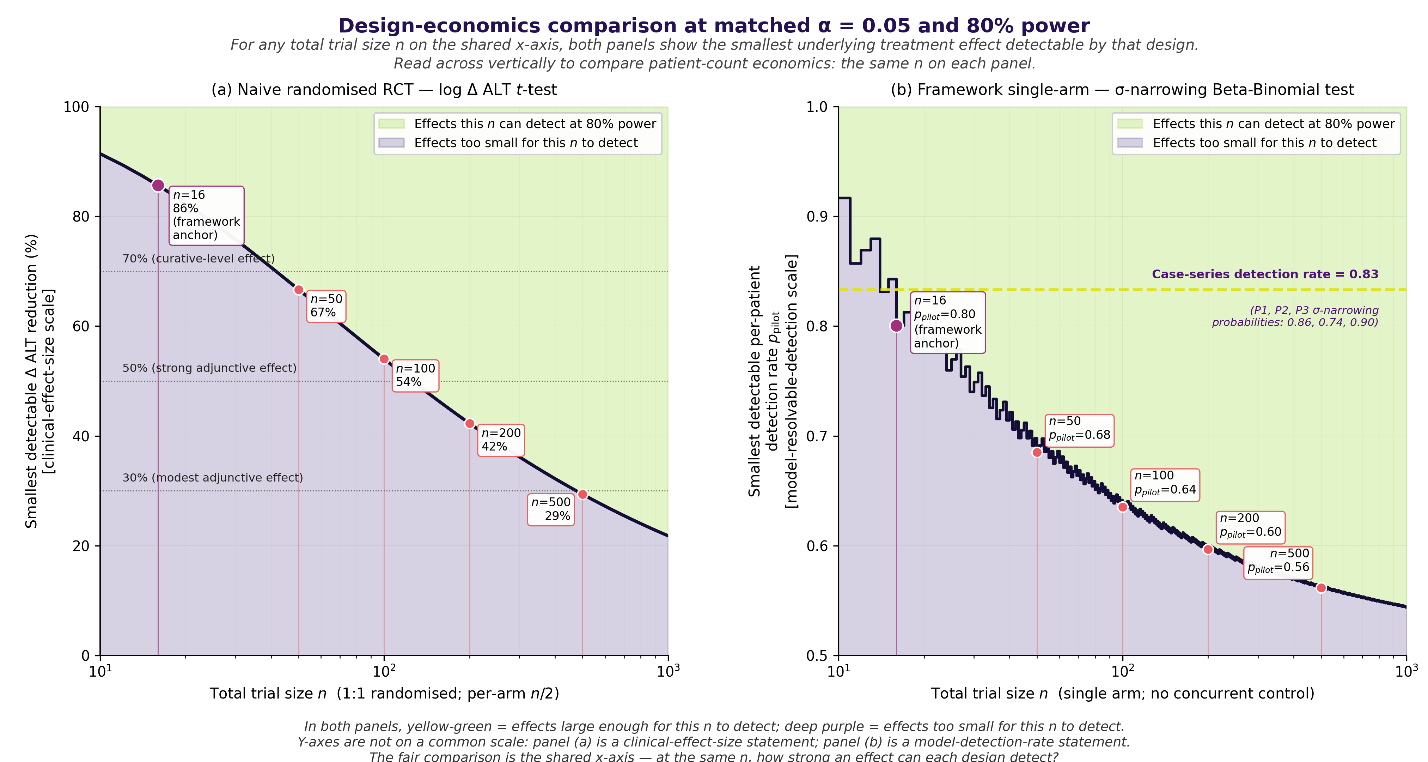

#### Supplementary Analysis 6. Severity-confound sensitivity: low cohort R² is not the same as no adjustment

This analysis demonstrates that the σ cohort regression with R² = 0.078 is doing severity adjustment. The cohort covariate is log peak ALT.

log peak ALT correlates with log σ in the SLTU sub-cohort at r = −0.32 (R² = 0.11 in all SLTU controls, n = 96; r = −0.51, R² = 0.26 in the well-identified pre-and-post-peak subset, n = 32). The correlation's sign is negative: in this cohort, bigger injuries have shorter σ. The M2 multivariable regression (log T ~ log λ + log peak ALT + log σ) uses log λ and log σ, and conditioning on those produces a useful predictive distribution that the fomepizole patients sit detectably below. λ shows a positive marginal correlation with peak ALT (r = +0.36, R² = 0.13): higher peak ALT correlates with faster clearance.

##### Supplementary Figure 18. Empirical peak-ALT correlations in the SLTU sub-cohort

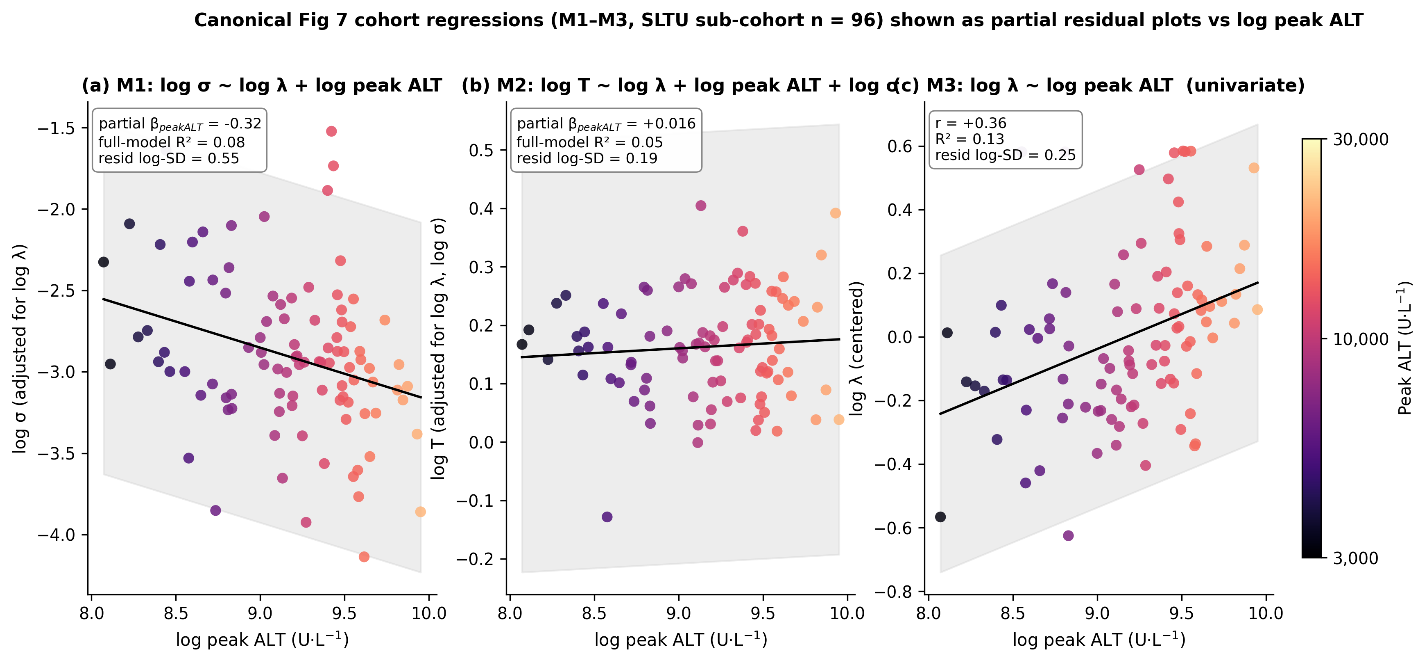

### Supplementary Tables

#### Supplementary Table 1. Mouse ALT and histological measurements

Per-mouse serial ALT measurements, peak ALT, time of peak ALT, ALT at cull, Gaussian ALT AUC computed to peak and to cull, and histological necrosis (fraction necrotic area on H&E-stained sections, InForm segmentation). n = 44 mice (44 paracetamol-treated; ALT AUC-necrosis regression cohort). Fasted-only controls (n = 5) excluded from this table; they showed no change in ALT from baseline (paired t-test, p = 0.602) and 0% histological necrosis.

| Mouse ID | Cull time (h) | Peak ALT (U/L) | Time of peak ALT (h) | ALT at cull (U/L) | ALT AUC to peak | ALT AUC to cull | Histological necrosis (fraction) |
| --- | --- | --- | --- | --- | --- | --- | --- |
| 206 | 8 | 10360 | 8 | 10360 | 42160 | 42160 | 0.49 |
| 207 | 8 | 9100 | 8 | 9100 | 37520 | 37520 | 0.334 |
| 209 | 8 | 9920 | 8 | 9920 | 40640 | 40640 | 0.336 |
| 210 | 8 | 7960 | 8 | 7960 | 32960 | 32960 | 0.23 |
| 211 | 16 | 11580 | 8 | 4540 | 47280 | 111760 | 0.248 |
| 212 | 16 | 12540 | 8 | 11220 | 50800 | 145840 | 0.441 |
| 213 | 16 | 8320 | 8 | 5180 | 33760 | 87760 | 0.157 |
| 214 | 16 | 7680 | 8 | 3620 | 31360 | 76560 | 0.349 |
| 215 | 14 | 11420 | 8 | 7920 | 46480 | 104500 | 0.415 |
| 216 | 24 | 5300 | 8 | 3640 | 21840 | 87840 | 0.341 |
| 217 | 24 | 8620 | 8 | 5740 | 35200 | 144320 | 0.339 |
| 218 | 24 | 8060 | 16 | 7940 | 87760 | 151760 | 0.517 |
| 219 | 24 | 8280 | 8 | 3600 | 33920 | 134560 | 0.316 |
| 220 | 24 | 8560 | 8 | 3820 | 34640 | 134720 | 0.382 |
| 221 | 16 | 13060 | 8 | 9220 | 53120 | 142240 | 0.397 |
| 222 | 8 | 12060 | 8 | 12060 | 49120 | 49120 | 0.256 |
| 223 | 36 | 11820 | 24 | 5720 | 194480 | 299720 | 0.591 |
| 224 | 36 | 6000 | 8 | 3640 | 24720 | 132480 | 0.314 |
| 225 | 36 | 8920 | 24 | 2100 | 95520 | 161640 | 0.19 |
| 226 | 11 | 13340 | 8 | 10360 | 54160 | 89710 | 0.314 |
| 227 | 48 | 13560 | 16 | 2440 | 93360 | 369640 | 0.324 |
| 228 | 48 | 13400 | 16 | 3160 | 125520 | 418440 | 0.551 |
| 229 | 42 | 14540 | 16 | 7740 | 136320 | 401920 | 0.484 |
| 230 | 48 | 14180 | 16 | 3500 | 145360 | 479880 | 0.335 |
| 231 | 48 | 16800 | 16 | 3940 | 169440 | 502480 | 0.364 |
| 232 | 13 | 12720 | 8 | 4500 | 51760 | 94810 | 0.232 |
| 233 | 24 | 13860 | 16 | 11340 | 125680 | 226480 | 0.557 |
| 234 | 13 | 13000 | 8 | 6140 | 53120 | 100970 | 0.26 |
| 235 | 48 | 12760 | 16 | 2380 | 125280 | 387920 | 0.538 |
| 236 | 48 | 8300 | 8 | 2320 | 33600 | 274760 | 0.548 |
| 237 | 48 | 8820 | 8 | 860 | 36080 | 199960 | 0.18 |
| 238 | 14 | 11300 | 8 | 9640 | 46160 | 108980 | 0.282 |
| 239 | 48 | 5960 | 24 | 740 | 94640 | 176120 | 0.215 |
| 240 | 31 | 14680 | 16 | 8540 | 128320 | 308730 | 0.345 |
| 241 | 48 | 8400 | 8 | 2280 | 34480 | 271680 | 0.431 |
| 242 | 16 | 13800 | 8 | 8480 | 55920 | 145040 | 0.32 |
| 243 | 48 | 8980 | 8 | 1680 | 36640 | 273720 | 0.399 |
| 244 | 48 | 17800 | 24 | 2160 | 163360 | 380800 | 0.467 |
| 245 | 48 | 6460 | 36 | 1380 | 174160 | 221200 | 0.311 |
| 246 | 48 | 6080 | 8 | 1020 | 25120 | 182320 | 0.216 |
| 247 | 48 | 12040 | 24 | 1060 | 109760 | 247640 | 0.239 |
| 248 | 48 | 5900 | 8 | 760 | 24240 | 140560 | 0.179 |
| 249 | 48 | 7620 | 8 | 720 | 31840 | 197000 | 0.223 |
| 250 | 48 | 8520 | 24 | 2160 | 162640 | 303760 | 0.51 |

#### Supplementary Table 2. Per-timepoint ALT-feature regression statistics (R²) in the murine cohort

Per-cull-timepoint summary of regression performance for four candidate ALT-trajectory features as predictors of histological necrosis in the murine cohort. n is the number of mice in each cull-time group. Values are R² for through-origin regression of percentage centrilobular necrosis on the corresponding ALT feature, demonstrating that ALT_AUC_to_cull maintained the strongest association with necrosis across the experimental period.

| Timepoint | n | ALT_at_cull | ALT_AUC_to_cull | Peak_ALT | ALT_AUC_to_peak |
| --- | --- | --- | --- | --- | --- |
| 8 | 5 | 0.924 | 0.924 | 0.924 | 0.924 |
| 13 | 5 | 0.935 | 0.960 | 0.941 | 0.940 |
| 16 | 6 | 0.928 | 0.944 | 0.935 | 0.935 |
| 24 | 6 | 0.938 | 0.971 | 0.958 | 0.866 |
| 36 | 5 | 0.879 | 0.927 | 0.899 | 0.911 |
| 48 | 17 | 0.882 | 0.907 | 0.869 | 0.756 |

#### Supplementary Table 3. Characteristics of the historical-control cohort and analysis subsets.

| **Characteristic** | **195 (analysed)** | **183 (FPCA)** | **130 (power calc.)†** |
| --- | --- | --- | --- |
| **Patients, n** | 195 | 183 | 130 |
| **Age, years** | 36 [24–44] | 36 [24–45] | 36 [26–44] |
| **Female sex, % (n)** | 51.8% (101) | 51.4% (94) | 47.7% (62) |
| **In-hospital mortality, % (n)** | 10.8% (21) | 10.4% (19) | 9.2% (12) |
| **Peak ALT, U/L** | 9,474 [6,467–12,483] | 9,246 [6,467–12,246] | 9,887 [7,147–13,150] |
| **Peak bilirubin, µmol/L** | 129 [70–208] | 129 [66–206] | 152 [92–246] |
| **Peak INR‡** | 4.9 [3.1–7.6] | 4.8 [3.0–7.6] | 5.1 [3.4–8.0] |
| **Peak lactate, mmol/L‡** | 3.2 [1.8–5.2] | 3.0 [1.8–5.0] | 3.3 [1.9–5.3] |

Continuous variables are median [IQR]; categorical variables are % (n). 183 (FPCA) = the analysed cohort minus 12 PC1 data-quality outliers.

† 130 (power pool) = patients with ≥ 1 ALT observation after the simulated treatment time. ‡ Peak INR available for 173 / 163 / 115 and peak lactate for 134 / 124 / 94 patients (195 / 183 / 130); remaining gaps are records with a prothrombin time but no INR, and patients in whom lactate was not measured.

#### Supplementary Table 4. 10-fold cross-validation residual summary vs full-data fit (n = 2,036 observations).

Predictive residuals under a 10-fold cross-validation scheme in which cohort-informed priors were derived from the training folds, compared with the full-data fit using the pre-specified weakly-informative priors (n = 2,036 observations). Cross-validation did not improve predictive accuracy, supporting use of the full-data fit.

| **Metric** | **Full-data fit** | **10-fold CV** |
| --- | --- | --- |
| R^2^ | 0.982 | 0.978 |
| RMSE (raw ALT, U/L) | 525.7 | 620.2 |
| MAE (raw ALT, U/L) | 329.7 | 406.0 |
| Mean relative residual | -0.111 | -0.174 |

#### Supplementary Table 5. Fomepizole cohort-regression statistics and per-patient mechanism predictions

| EMG parameter | Mechanistic prediction | Test type | P1 | P2 | P3 | P4 | BB composite mean (95% CrI) |
| --- | --- | --- | --- | --- | --- | --- | --- |
| σ (injury duration) | shorter than expected given peak ALT | one-sided | 0.86 | 0.74 | 0.90 | 0.34 | 0.64 [0.18, 0.98] |
| T (peak timing) | earlier than expected given peak ALT | one-sided | 0.78 | 0.79 | 0.73 | 0.39 | 0.62 [0.15, 0.97] |
| λ (clearance) | unchanged (no systematic deviation) | two-sided equivalence | 0.45 | 0.55 | 0.65 | 0.02 | no composite (see Supp Fig 12) |

#### Supplementary Table 6. Bayesian convergence diagnostics across all 195 patients (4 chains × 2,000 draws).

188 of 195 patients met R̂ ≤ 1.01 on all parameters; 7 exceeded 1.01 (5 exceeded 1.05); median R̂ across patients and parameters was 1.001. The total divergence count across the cohort was 313. Trace plots and posteriors for the 7 patients exceeding R̂ = 1.01 are shown in Supplementary Figure 19. Five patients with divergence (17, 36, 86, 134, 147) had data sparsity (n=3 to 4 observations to fit) and one further patient (163) had limited post-peak data (n=2 post-peak observations).

| **Parameter** | **R-hat median** | **R-hat max** | **ESS bulk median** | **ESS tail median** |
| --- | --- | --- | --- | --- |
| A (peak release rate) | 1.0011 | 1.94 | 3,005 | 3,268 |
| σ (peak width) | 1.0011 | 2.16 | 2,879 | 3,329 |
| T (peak time offset) | 1.0010 | 2.24 | 3,295 | 3,518 |
| λ (decay time) | 1.0008 | 2.23 | 4,431 | 4,145 |
| ingestion time | 1.0011 | 2.12 | 3,166 | 3,463 |
| measurement noise | 1.0007 | 1.82 | 5,086 | 4,575 |

##### Supplementary Figure 19. Convergence diagnostics for the seven patients with R̂ > 1.01 on any parameter.

For each historic-cohort patient whose maximum split-R̂ exceeded 1.01, the four MCMC chains (left) and the per-chain posterior density (right) are shown for the parameter with the highest R̂. These patients are retained in the cohort.

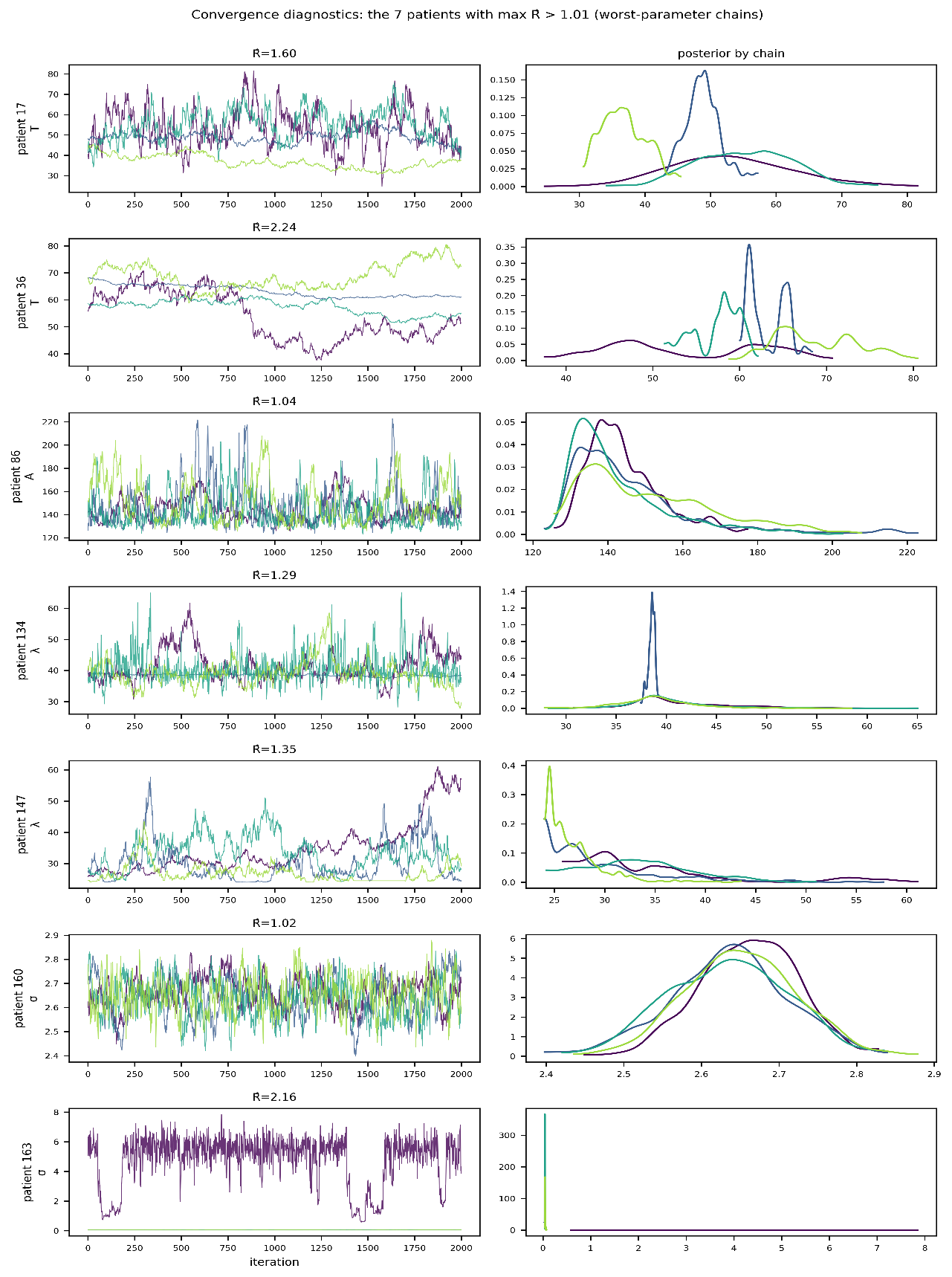

### Supplementary Methods (expanded)

#### SM1. Preclinical APAP murine model - full procedure

Fifty twelve-week-old male C57BL/6J wild-type mice (Charles River Laboratories, UK) were housed in groups of five on a 12-hour light/dark cycle. Mice were allocated to cull-timepoint groups sequentially (not randomised); blinding to group allocation was not possible given the serial-sacrifice design. The study was exploratory and no formal sample-size calculation was performed. One week prior to APAP administration, baseline plasma ALT was quantified from a 40-microlitre tail-vein bleed (plasma separated by centrifugation at 2,000 g for 10 minutes). At 14 h prior to APAP administration, food was removed in keeping with the published protocol of Starkey Lewis et al. At t = 0, n=45 mice received a single paracetamol injection (350 mg/kg intraperitoneal in warm sterile saline); n=5 fasted-only control mice were culled at t = 0 without receiving APAP to confirm that baseline ALT values were valid (paired t-test on ALT change from baseline, p = 0.602; 0% histological necrosis). Standard chow and wet mash were returned post-administration and mice transferred to a warming cabinet (28 degrees C). Serial tail-vein blood samples (40 microlitres) were collected at 8, 16, 24, 36 and 48 h post-APAP until each mouse reached its scheduled or humane-endpoint cull; a terminal sample was obtained by cardiac puncture at cull. A phenotypic humane endpoint scoring system was applied throughout (per Starkey Lewis et al.). Seven mice required humane cull outside pre-specified timepoints (one at 11 h, two at 13 h, two at 14 h, one at 31 h, one at 42 h); they were retained in the analysis at the median grouping (11/13/14 h grouped at 13 h; 31/42 h within the 36-h group). One mouse was found dead at 7 h post-injury with no clear cause on necropsy and was excluded. Final group sizes: 0 h (fasted-only controls) n = 5; 8 h n = 5; 13 h n = 5; 16 h n = 6; 24 h n = 6; 36 h n = 5; 48 h n = 17 (enriched to support secondary experimental aims). Total n = 49 (44 APAP-treated mice contributed to the ALT AUC-necrosis regression). Animal studies were approved by the University of Edinburgh Animal Welfare and Ethical Review Body (093-LFR-24); the study was not pre-registered.

Histology and ALT assay. At cull, livers were fixed in 10% neutral buffered formalin for 8 h, dehydrated with 70% ethanol, embedded in paraffin wax, and sectioned at 4 micrometres. Sections were stained with haematoxylin and eosin (H&E) using standard protocols. ALT was measured by a blinded laboratory service using the method of Bergmeyer et al. Intra-assay precision CV < 4%; inter-assay precision CV < 8%. Tissue necrosis was quantified as percentage necrotic area across a minimum of 10 digitally defined regions of interest per mouse, delineated using InForm tissue segmentation software (Akoya Biosciences). InForm defined subset regions excluding vessel lumina and air spaces; percentage necrotic area was quantified within these regions by automated classification of H&E-stained tissue.

#### SM2. Bayesian EMG model - full specification

ALT(t) is modelled as the convolution of a Gaussian injury process (a transient hepatocyte-release rate centred at time T with width σ and amplitude A) with a first-order clearance process with time constant λ. Convolving G(s) = A·exp(−(s−T)²/(2σ²)) with the clearance impulse response (1/λ)·exp(−(t−s)/λ) for s ≤ t yields the closed-form exponentially-modified Gaussian: ALT(t) = y₀ + A·σ·√(π/2)·exp(½(σ/λ)² + (T−t)/λ)·erfc((λ(T−t)+σ²)/(λ·σ·√2)). The five structural parameters are peak release rate A, injury duration σ, time-to-peak T, clearance time constant λ, and baseline y₀; y₀ is fixed at 30 U·L⁻¹ (within the normal range) and the other four are inferred per patient. The model additionally infers each patient's ingestion time and an absolute measurement-noise scale σ_meas.

Observations were modelled with absolute (homoscedastic) Gaussian error: ALT_obs(t) ~ Normal(ALT(t), σ_meas), with σ_meas an inferred parameter. Measurement error was modelled as absolute (homoscedastic) Gaussian; reflecting the approximately constant absolute analytical precision of the ALT assay across its range.

Priors were weakly informative and centred on values consistent with published APAP-DILI literature: A ~ Exponential(λ = 1/500); σ ~ Gamma(μ = 14, σ = 7); T ~ Gamma(μ = 60, σ = 12); λ ~ TruncatedNormal(50, 12; lower = 24, upper = 86); σ_meas ~ TruncatedNormal(4.576, 4.576; lower = 0.1, upper = 704). The ingestion-time prior differs by sub-cohort: for patients with a documented last-ingestion time (SLTU sub-cohort, n = 96), ingestion time ~ TruncatedNormal(0, 12; lower = −168, upper = first sample time); for patients without documented ingestion (NHS Lothian sub-cohort, n = 99), ingestion time ~ TruncatedNormal(−51.83, 19.71; upper = −4) h, derived from the median post-overdose presentation time in the SLTU sub-cohort.

Inference used PyMC v5.21.1, NUTS sampler, 4 chains × 2,000 sampling iterations with 1,000 tuning iterations each, target_accept = 0.99. Convergence diagnostics are summarised in Supplementary Table 5

#### SM3. PACE-fPCA - full specification

PACE-fPCA on residual trajectories. Functional principal component analysis was performed using PACE (Principal Analysis by Conditional Expectation; Yao et al. 2005) on the log₂ residuals r(t) = log₂(ALT_observed(t) / ALT_predicted(t)) evaluated at observed timepoints, where ALT_predicted is the posterior-mean EMG curve from a counterfactual (‘predict-mode’) fit in which each patient's post-cutoff observations are held out during fitting (distinct from the full-data SM2 fits used for the cohort parameter posteriors and the fomepizole comparison). PACE smooths the cohort mean function and the covariance surface separately (fixed smoothing bandwidths), then projects each patient's sparse trajectory onto the smoothed eigenbasis via conditional expectation under the cohort covariance, allowing the method to score patients with as few as three observations without bias from sparse individual coverage. Three components were retained, capturing 95.0% of residual variance (PC1: 86.3%, PC2: 6.5%, PC3: 2.2%; canonical run residual_fpca_full_vs_predict.ipynb). PC1 carries the dominant injury-duration / clearance-modification signal.

#### SM4. LOO regression - full derivation

PC1_adj is constructed by regressing PC1 on log(λ) by leave-one-out (LOO) regression. For each patient i, the regression is re-fitted on the other n - 1 patients and PC1_adj,i = PC1_i - (intercept + β · log(λ_i)). The full-cohort coefficients are intercept = 86.34 and β = -23.57; full-data R² = 0.53; LOO variance reduction = 0.52; n = 183. Single-predictor (log(λ) alone) was selected by LOO model comparison; alternative two-predictor candidates that included log(σ_inj) or ingestion time were tested but did not meet the LOO improvement threshold.

#### SM5. Power analysis - full procedure

Treatment effects were simulated by accelerating each historic-cohort patient's ALT clearance by a fraction δ (scaling the posterior clearance rate by 1 + δ). Five dose cohorts were defined at clearance-increase fractions of 10%, 20%, 30%, 40% and 50% of an overall effect-size multiplier m, swept over m ∈ {0, 0.2, 0.4, 0.6, 0.8, 1.0, 1.2, 1.5, 2.0}; at m = 1 the cohorts correspond to 10–50% clearance increases, and at the maximum m = 2 the top cohort corresponds to a 100% increase.

For each m, 2,000 bootstrap trials were assembled by sampling 15 patients without replacement from the 130-patient eligible pool and assigning 3 to each of the five dose cohorts; the remaining 115 patients served as an untreated dose-zero anchor. Two analyses were scored on the same simulated treated patients: (i) the framework - a permutation slope test of PC1_adj on dose using the 115 dose-zero patients together with the 15 treated patients (299 permutations per trial); and (ii) the planned analysis of the MAIL trial(15), a published exemplar of current phase-1 dose-escalation practice - a Jonckheere–Terpstra ordered-alternatives test on the per-patient anchored log-ALT decay slope across the five treated cohorts (15 treated patients only). Both used a one-sided α = 0.05.

False-positive rates were verified under the global null (m = 0): 5.7% for the framework and 4.2% for the protocol analysis, both within the nominal 5% to bootstrap precision. The minimum detectable effect at 80% power was obtained by linear interpolation between adjacent m values: m* = 0.489 for the framework (top-cohort clearance increase ≈ 24.5%) and m* = 1.349 for the protocol analysis (≈ 67.5%), a 2.76× reduction.

#### SM6. Cohort-regression-adjusted comparison for fomepizole - full procedure

Three log-scale OLS regressions were fitted on the SLTU sub-cohort posterior parameters: log σ ~ log λ + log peak ALT (M1); log T ~ log λ + log peak ALT + log σ (M2); and log λ ~ log peak ALT (M3). To propagate control fit-uncertainty (errors-in-variables, EIV) the regressions were re-fitted K = 1000 times: in each replicate, one posterior sample per control patient was drawn at random, and the three OLS regressions were re-fitted. Each fomepizole patient's cohort prediction was then formed per MCMC draw from a randomly chosen replicate, composing control fit-uncertainty (between replicates), regression-coefficient uncertainty (within replicate, via Cholesky of Cov(β̂)), and residual scatter (ε ~ Normal(0, σ_resid²)). For per-patient inference, P(σ_fome < σ_cohort) was computed as the posterior probability that the patient's posterior draw of log σ lay below the cohort predictive distribution conditional on the patient's own λ and peak ALT (and for T, additionally σ). The across-patient Beta-Binomial composite uses the exact mixture posterior π | data ∝ Σ_y∈{0,1}^n [Π_i p_i^{y_i} (1 - p_i)^{1 - y_i}] · Beta(1 + Σy, 1 + n - Σy), enumerated exactly for n = 4 (16 latent success patterns). Fixed random seed = 42 throughout. Severity-confound sensitivity (Supplementary Analysis 6) re-computes per-patient probabilities under a peak-ALT-restricted non-parametric comparator and confirms the regression-based adjustment matches a direct severity-matched comparison to within 0.01 for P1 and P3.

The cohort regressions are not fit on per-control posterior means; instead, one posterior sample per control patient is drawn at random, the three log-scale OLS regressions are fit on the resulting per-control values, and this is repeated K = 1,000 times to form K replicate regression coefficient sets. Each fomepizole patient's cohort prediction is then formed per MCMC draw by sampling a replicate at random, drawing β from N(β̂_replicate, Cov(β̂_replicate)) via Cholesky, and adding residual scatter ε ~ N(0, σ̂_resid²). This composes three uncertainty sources: between-replicate (control-fit), within-replicate (regression-coefficient), and residual. The across-patient Beta-Binomial composite uses the exact mixture posterior π | data ∝ Σ_y∈{0,1}^n [Π_i p_i^{y_i} (1 − p_i)^{1−y_i}] · Beta(1 + Σy, 1 + n − Σy), enumerated exactly for n = 4 (16 latent success patterns; no Monte Carlo).
